# Association of Inflammatory Burden Index with All-Cause and Cardiovascular Mortality Among Individuals with CKM Syndrome

**DOI:** 10.64898/2025.12.10.25342023

**Authors:** Yuqi Qin, Luoning Gou

**Affiliations:** Tongji Hospital, Tongji Medical College, Huazhong University of Science and Technology, Wuhan 430030, China; Department of Endocrinology, Tongji Hospital, Tongji Medical College, Huazhong University of Science and Technology, Wuhan 430030, China; Hubei Clinical Medical Research Center for Endocrinology and Metabolic Diseases, Hubei, China; Branch of National Clinical Research Center for Metabolic Diseases, Hubei, China

**Keywords:** Inflammatory Burden Index, CKM, All-cause mortality, Cardiovascular mortality, Inflammation

## Abstract

**Background:** Cardiovascular-Kidney-Metabolic (CKM) syndrome, as defined by the American Heart Association (AHA) in 2023, encompasses pathophysiological interconnections among the cardiovascular, renal, and metabolic systems, which collectively lead to multi-organ dysfunction. The Inflammatory Burden Index (IBI), a newly developed inflammatory marker, has an unclarified association with mortality in the con text of CKM syndrome. Our study aimed to explore the relationships between IBI and mortality in this pop ulation, with the goal of facilitating early clinical intervention.

**Methods:** 6,091 NHANES (1999-2018) participants with CKM syndrome were analyzed. Weighted Cox models and Restricted cubic spline (RCS) analysis assessed IBI’s association with all-cause and cardiovasc ular mortality. We tested proportional hazards (Schoenfeld residuals), model performance (ROC curves), a nd confounding (subgroup/stratified analyses).

**Results:** IBI independently predicted mortality after full adjustment: All-cause mortality: HR=1.13, 95% C I:1.05–1.21, *p*=0.002;Cardiovascular mortality: HR=1.35, 95% CI:1.21–1.51, *p*<0.001.The fully adjusted model (AUC>0.8) showed optimal predictive performance. IBI exhibited non-linear (“U-shaped” all-cause, “J-shaped” cardiovascular) mortality relationships. The highest IBI quartile (Q4) robustly predicted cardiovascular death (HR=1.77, 95% CI:1.18–2.67).Subgroup and stratified analyses revealed that male s ex, advanced age, smoking, and high-risk chronic kidney disease (CKD) were synergistic factors exacerbating the association between IBI and all-cause mortality risk, while female sex, smoking, diabetes, and meta bolic abnormalities were synergistic factors exacerbating the association with cardiovascular mortality risk.

**Conclusion:** Our study is the first to demonstrate that IBI exhibits significant non-linear associations with both all-cause and cardiovascular mortality in patients with CKM syndrome and serves as a risk factor for these patients. Monitoring the Inflammatory Burden Index holds important clinical significance for assessing mortality risk in individuals with CKM syndrome.

## 1. Introduction

Type 2 diabetes mellitus (T2DM), cardiovascular disorders (CVD), and chronic kidney impairment (CKD) currently represent three predominant pathological conditions that pose significant threats to global public health. These non-communicable diseases have emerged as leading causes of morbidity and mortality worldwide. Substantial evidence shows that these three conditions frequently co-occur^1^. To better understand the intricate relationships between these conditions, the American Heart Association (AHA) formally proposed the unifying concept of Cardiovascular-Kidney-Metabolic (CKM) syndrome in October 2023. This novel classification underscores the profound interdependencies and pathophysiological interplay among metabolic dysfunction, cardiovascular disease, and renal impairment. CKM syndrome is defined as health impairments attributable to the connections among obesity, diabetes, CKD, and CVD, including heart failure, atrial fibrillation, coronary heart disease, stroke, and peripheral artery disease^2^. A research project using data from the China Health and Retirement Longitudinal Study (CHARLS) has reported that CKM syndrome represents a notable risk factor for disability in activities of daily living (ADL) among middle-aged and elderly individuals. The risk of mobility decline and disability increased significantly with advancing CKM stages^3^. Another prospective cohort study found that higher CKM syndrome stages were associated with greater risks of adverse events following non-cardiac surgery. Specifically, CKM stage ≥3 was significantly associated with major adverse cardiovascular events (MACE), all-cause mortality, and non-MACE complications within 30 days postoperatively^4^. Suboptimal CKM health is a key contributor to all-cause mortality, imposing a substantial burden on global public health systems and socioeconomic resources^5^. Hence, elucidating the risk factors for CKM syndrome to enable early prediction and intervention is crucial for preventing adverse outcomes in affected individuals.

A substantial body of research has confirmed that inflammatory processes occupy a critical position in the pathological mechanisms underlying cardiovascular diseases, renal diseases, and metabolic disturbances. The inflammatory pathways involved in coronary atherosclerosis include endothelial cell migration leading to intimal thickening^6^, monocyte recruitment and differentiation into macrophages^7^, T-lymphocyte activation of monocyte-macrophages to produce pro-inflammatory and anti-inflammatory cytokines, and the amplification of local inflammatory responses by inflammatory factors^8,9^. Macrophages degrade oxidized low-density lipoprotein cholesterol, forming lipid-rich necrotic cores that stimulate vascular smooth muscle cell migration and collagen cap formation, resulting in stable fibroatheromas^10^. Under chronic inflammation, macrophage activity thins the fibrous cap, leading to thin-cap fibroatheromas (TCFAs) and the formation of vulnerable plaques prone to rupture and thrombosis^11,12^. Moreover, chronic kidney disease (CKD) is linked to increased concentrations of inflammatory markers. Experimental studies on animals have shown that the activation of the NLRP3 (NOD-like receptor family pyrin domain containing 3) inflammasome in renal parenchymal cells may induce diabetic kidney disease (DKD). In contrast, the suppression of IL-1 receptor signaling or mitochondrial oxidative stress can alleviate DKD^13^. Free fatty acids (FFAs) have been demonstrated to trigger metabolic inflammation and insulin resistance through at least four distinct mechanisms: i) activation of endoplasmic reticulum (ER) stress^14^, ii) stimulation of Toll-like receptor 4 (TLR4) signaling^15^, iii) activation of protein kinase C theta (PKCθ)/PKC delta (PKCδ)^16^, and iv) the activation of protein kinase R (PKR)^17^. Cytokines including tumor necrosis factor-alpha (TNF-α), interleukin-1 beta (IL-1β), and interleukin-6 (IL-6) are capable of exacerbating metabolic inflammation. As a result, inflammatory responses are an essential component in the onset and progression of disorders related to CKM syndrome.

The Inflammatory Burden Index (IBI) is a novel biomarker designed to quantify the degree of inflammation, offering significant clinical applicability. The proposal was first presented based on findings indicating that the levels of pro-inflammatory cytokines (such as C-reactive protein, CRP) and the presence of inflammatory cells (including neutrophils and lymphocytes) are heightened in the synovial tissue, synovial fluid, and peripheral blood of patients with osteoarthritis^18–20^. Xie et al. first developed IBI in a multicenter study involving 6,359 patients with various cancers to assess inflammatory burden and predict cancer prognosis. Their findings revealed a significant non-linear association between IBI and cancer patient survival, confirming IBI as a feasible and promising prognostic biomarker in oncology^21^. A multicenter cohort study conducted in China, which included 295 patients with acute ischemic stroke who underwent endovascular thrombectomy, found that elevated IBI levels were found to be positively correlated with a heightened risk of unfavorable outcomes within 90 days^22^. Despite the increasing identification of inflammatory markers indicating severity and predicting prognosis in CKM syndrome, the optimal inflammatory index for comprehensively evaluating inflammatory burden remains to be explored.

Thus, the present study drew upon data from the National Health and Nutrition Examination Survey (NHANES) database to explore the relationship between the Inflammatory Burden Index (IBI) and all-cause as well as cardiovascular mortality among individuals with CKM syndrome. We aimed to facilitate early prevention of CKM syndrome by establishing the correlation between IBI and mortality.

## 2. Methods

### 2.1 Study participants

The National Health and Nutrition Examination Survey (NHANES) is a nationwide investigation formulated to gather data concerning the nutritional and health conditions of the non-institutionalized civilian population in the United States. This study utilized de-identified data from the NHANES database. As such, it was exempt from institutional review board approval and the requirement for informed consent was waived. The study was conducted in accordance with the principles outlined in the Declaration of Helsinki.By utilizing a sophisticated, stratified, multistage probability sampling approach, NHANES ensures that its samples are nationally representative^23^. Detailed information about the study design and data of NHANES can be accessed via its official website (www.cdc.gov/nchs/nhanes/). This study utilized data from ten NHANES cycles spanning 1999 to 2018 (excluding the 2011-2014 cycle due to the absence of C-reactive protein data). Participants who were pregnant or for whom the 10-year cardiovascular disease (CVD) risk could not be calculated using the basic Predicting Risk of CVD Events (PREVENT) equations were initially excluded ^24^. The exclusion criteria, outlined in Figure 1, detail the value ranges of variables incompatible with the basic PREVENT equations. Additionally, we removed participants with inapplicable follow-up data or missing values for covariates including statin use and the poverty-income ratio (PIR). Finally, participants lacking data for the Inflammatory Burden Index (IBI) or its component indices were excluded (Figure 1). The ultimate analytical sample consisted of 6,091 participants ranging in age from 30 to 79 years old. The cohort size consists of 6091 participants who were followed for a median of 10.4 years, with the follow - up time ranging from 0.2 to 20.8 years.

**Figure 1.**
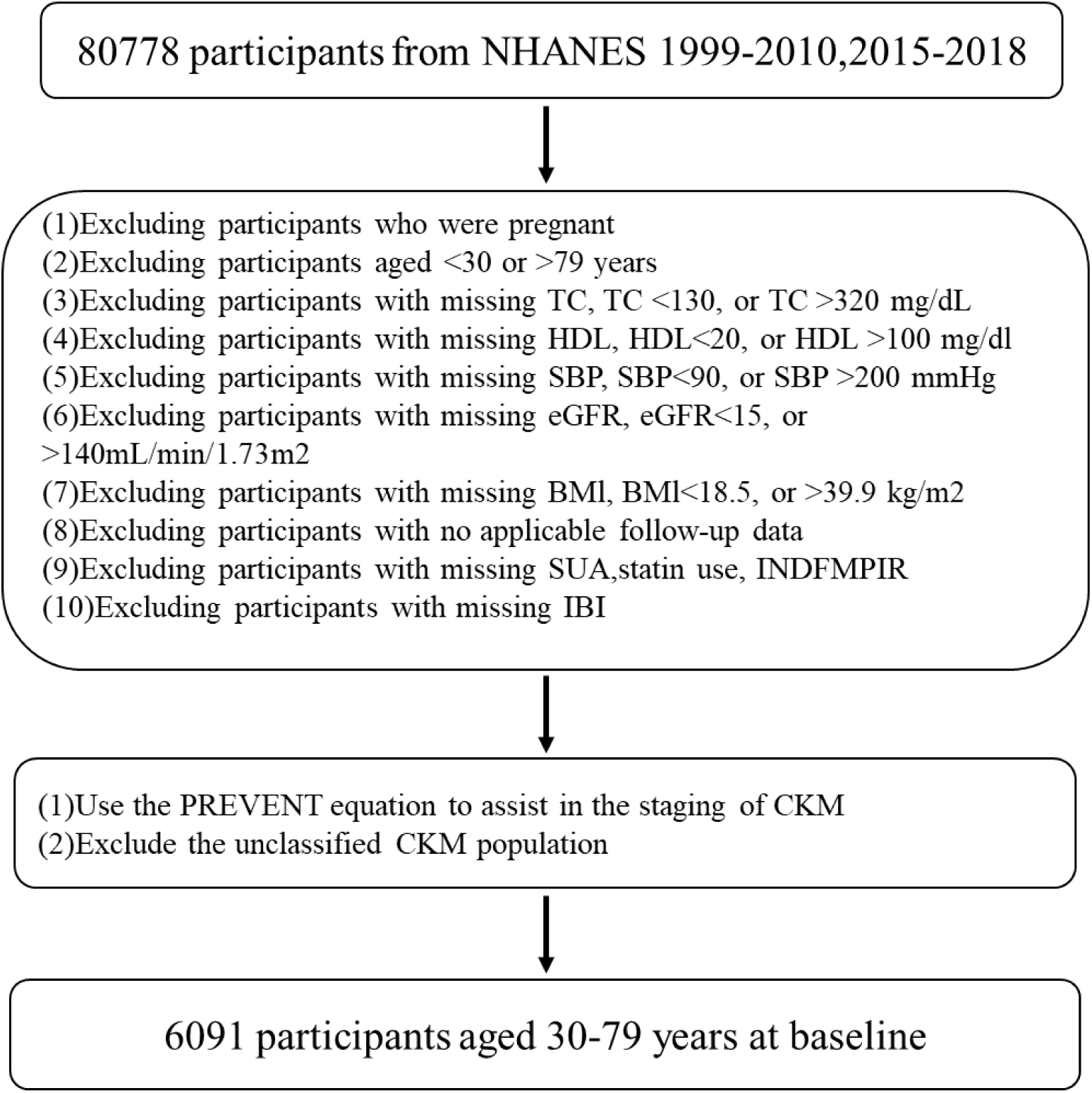
Flowchart of the study population.

### 2.2 Definition of the inflammation burden index

The Inflammatory Burden Index (IBI) was derived from three biomarkers: neutrophil count, lymphocyte count, and C-reactive protein (CRP). It was calculated using the following formula:

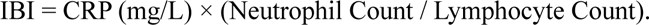

A higher Inflammatory Burden Index (IBI) score signifies a more severe inflammatory burden, while a lower score indicates a less pronounced inflammatory burden.

### 2.3 Definition of CKM stage

CKM syndrome includes individuals who are at risk of developing cardiovascular disease (CVD) as well as those with pre-existing CVD^2^. The stages are defined as follows:

CKM Stage 0: Individuals without overweight/obesity, metabolic risk factors (hypertriglyceridemia, hypertension, diabetes, metabolic syndrome), chronic kidney disease (CKD), or subclinical/clinical CVD. CKM Stage 1: Individuals with overweight/obesity, abdominal obesity, or adipose tissue dysfunction, but without other metabolic risk factors, CKD, or subclinical/clinical CVD. This stage is characterized by:

1. Body mass index (BMI) ≥25 kg/m²;
2. Waist circumference (WC) is ≥102 cm for males or ≥88 cm for females;
3. Fasting blood glucose (FBG) concentrations range from 100 to 124 mg/dL, or hemoglobin A1c (HbA1c) levels are in the range of 5.7% to 6.4%.

CKM Stage 2: Individuals with metabolic risk factors (including hypertriglyceridemia, hypertension, diabetes mellitus, and metabolic syndrome) or those in moderate-to-high-risk stages of chronic kidney disease (CKD) (as specified by the KDIGO criteria based on estimated glomerular filtration rate [eGFR] and urine albumin-to-creatinine ratio [UACR]). The eGFR was computed using the 2021 Chronic Kidney Disease Epidemiology Collaboration (CKD-EPI) creatinine equation, which excludes coefficients related to race and ethnicity^25^.

CKM Stage 3: Individuals with subclinical CVD or its risk equivalents: high predicted 10-year CVD risk or very-high-risk KDIGO CKD stage.

1. High 10-year CVD risk was defined as a risk ≥20%, determined using the basic Predicting Risk of CVD Events (PREVENT) equations.
2. Very-high-risk CKD per KDIGO classification was defined as UACR ≥300 mg/g, eGFR ≤45–59 mL/min/1.73 m², UACR ≥30 mg/g with eGFR ≤30–44 mL/min/1.73 m², or eGFR ≤29 mL/min/1.73 m². CKM Stage 4: Individuals with clinical CVD (self-reported diagnosis including heart failure, coronary heart disease, angina, myocardial infarction, or stroke).

### 2.4 Definition of clinical outcomes

Mortality data were retrieved from the NHANES Public-Use Linked Mortality File up to December 31, 2019. This file employed probabilistic matching methods to connect participants with the National Death Index (NDI) administered by the National Center for Health Statistics (NCHS). The causes of death were identified in accordance with the International Classification of Diseases, Tenth Revision (ICD-10), and the specified mortality outcomes underwent reclassification. All-cause mortality was defined as death resulting from any cause (ICD-10 codes: A00-Z99). Cardiovascular mortality included deaths caused by heart disease (ICD-10: I00-I09, I11, I13, I20-I51) and cerebrovascular disease (ICD-10: I60-I69).

### 2.5 Covariate assessment

We included various covariates potentially influencing outcomes. Age, gender, racial/ethnic background, family poverty-income ratio (PIR), smoking habits, alcohol consumption patterns, medication usage, and disease status were gathered through standardized questionnaires during household interviews. Body mass index (BMI), waist circumference (WC), and blood pressure (BP) were measured during physical examinations carried out in mobile examination units. Laboratory indicators included C-reactive protein (CRP), neutrophil count, lymphocyte count, fasting blood glucose (FBG), low-density lipoprotein cholesterol (LDL-C), high-density lipoprotein cholesterol (HDL-C), hemoglobin A1c (HbA1c), total cholesterol (TC), triglycerides (TG), urine albumin, creatinine (Cr), and estimated glomerular filtration rate (eGFR). Systolic blood pressure (SBP) and diastolic blood pressure (DBP) were determined as the average of three separate readings. Hypertension was defined as a systolic blood pressure of ≥130 mm Hg, a diastolic blood pressure of ≥ 80 mm Hg, a confirmed diagnosis by a physician, or current use of antihypertensive drugs^26^. Diabetes mellitus was defined as FBG level >126 mg/dL, HbA1c ≥6.5%, physician diagnosis, or current use of insulin or hypoglycemic agents^27^. Metabolic syndrome (MetS) was defined as the presence of ≥3 of the following criteria: WC ≥102 cm in men or ≥88 cm in women; HDL-C <40 mg/dL in men or <50 mg/dL in women; TG ≥150 mg/dL; Elevated BP (as defined above); FBG ≥100 mg/dL. Chronic kidney disease (CKD) was categorized into low-risk, moderate-to-high-risk, and very-high-risk groups according to KDIGO criteria based on eGFR and urine albumin-to-creatinine ratio (UACR)^28^. Alcohol consumption status was classified as follows: Heavy drinking: Consumption of ≥3 drinks per day for women or ≥4 drinks per day for men, or binge drinking (≥4 drinks for women or ≥5 drinks for men on the same occasion) on ≥5 days per month. Moderate drinking: Consumption of 2 drinks per day for women or 3 drinks per day for men, or binge drinking on ≥2 days per month. Light drinking: Consumption patterns not meeting criteria for heavy or moderate drinking. Never drinker: Lifetime consumption of <12 alcoholic drinks. Smoking status was defined as: Never smoker: Lifetime consumption of <100 cigarettes. Former smoker: Individuals who have consumed 100 or more cigarettes in their lifetime but are not smoking at present. Current smoker: Individuals who have consumed 100 or more cigarettes in their lifetime and were smoking at the time of the survey. For detailed information on the interpretation, access, and calculation of each covariate, please refer to the official NHANES Analytic Guidelines.

### 2.6 Statistical analysis

All statistical analyses were performed using R software version 4.4.3. A two-sided p < 0.05 was considered statistically significant. Survey weighting was utilized to address the complex multistage probability sampling framework of the NHANES dataset, thereby ensuring that the estimates for the U.S. population are both accurate and representative. Adjustments to sampling weights were made with consideration to selection probabilities and response rates. Specifically, weights were calculated as: wtmec (4-year) * 2/10 for the 1999-2002 cycles and wtmec (2-year) * 1/10 for the 2003-2018 cycles. Multiple imputation was performed for three covariates with >10% missing data (LDL-C, HbA1c, fasting glucose). The pattern of missing data and diagnostics for imputation are provided in Supplemental Figure 1. One randomly chosen imputed dataset was utilized for analytical purposes. The Inflammatory Burden Index (IBI) underwent log transformation (ln_IBI) and was assessed as a continuous variable using weighted Cox proportional hazards regression to examine its association with all-cause mortality in the CKM population. Three regression models were constructed: Model 1: Unadjusted. Model 2: Adjusted for demographic covariates (age, sex, race/ethnicity, poverty-income ratio (PIR)). Model 3: Fully adjusted for age, sex, race/ethnicity, PIR, BMI, eGFR, urine albumin-to-creatinine ratio (UACR), uric acid, SBP, DBP, HDL-C, LDL-C, HbA1c, smoking status, alcohol consumption, statin use, antihypertensive medication use, hypertension, diabetes, metabolic syndrome (MeTS), CKD, and CKM stage. Potential multicollinearity was evaluated via variance inflation factors (VIF). Only covariates with a VIF < 5 were retained in the final models. Comprehensive diagnostics for multicollinearity are presented in Supplemental Table S1. The proportional hazards assumption was examined using Schoenfeld residuals. Model performance was assessed and compared using the concordance index (C-index) and Wald tests. Receiver operating characteristic (ROC) curves were generated for all three models, and their predictive accuracy was compared using the area under the curve (AUC). The dose-response relationship between ln_IBI and all-cause mortality was characterized using generalized additive models (GAMs) with smoothing curve fitting (restricted cubic splines, RCS). The number of knots was determined by minimizing the Akaike Information Criterion (AIC). If a non-linear relationship was detected, the CKM population was stratified into low-IBI and high-IBI groups based on the inflection point. The baseline characteristics of these groups were compared and tabulated. For continuous variables, analysis was performed using survey-weighted linear regression, while survey-weighted chi-square tests were applied to categorical variables. To assess the impact of covariates on model effects and mitigate confounding, subgroup and stratified analyses were conducted according to age, sex, race, disease status (hypertension, diabetes, MeTS, CKM stage), and lifestyle factors (smoking, alcohol consumption). Participants were also categorized into quartiles (Q1-Q4) based on ln_IBI levels within each model. Odds ratios (ORs) and 95% confidence intervals (95% CIs) were calculated, with the Q1 group as the reference. The results were visualized through forest plots. Continuous variables are reported as weighted means (95% CI), and categorical variables as weighted percentages (95% CI). To confirm the stability of the association between IBI and all-cause mortality in the CKM population, a further subgroup analysis was conducted by stratifying participants according to their baseline characteristics (including age, sex, race/ethnicity, smoking status, alcohol consumption status, hypertension, diabetes, MetS, CKD risk, and CKM stage). Statistical significance for interaction terms was tested, and forest plots were employed for visualization purposes. Stratified analyses adjusting for age, sex, and race were further conducted to control for potential confounding in the association between IBI and mortality. The association between IBI and cardiovascular mortality was analyzed using methods analogous to those described above for all-cause mortality.

## 3 Results

### Baseline Characteristics of Participants Stratified by IBI Cut-Off Values

Table 1 demonstrates that CKM patients stratified by the Inflammatory Burden Index (IBI) cutoff value (determined by subsequent RCS analysis) exhibited significantly higher overall health risks in the high-IBI group compared to the low-IBI group. Specifically, the high-IBI group had an older age, more severe obesity and metabolic dysregulation (characterized by poor glycemic and lipid control), higher hypertension and cardiovascular risk, worse renal function (lower eGFR), greater chronic disease burden and behavioral risks (e.g., higher smoking prevalence), lower statin usage, more advanced disease staging (CKM stage), and higher mortality rates. Conversely, the low-IBI group exhibited the opposite characteristics. Thus, high IBI is strongly associated with adverse health profiles and mortality outcomes, supporting its utility as a comprehensive health risk indicator in the population with CKM syndrome.

**Table 1.**
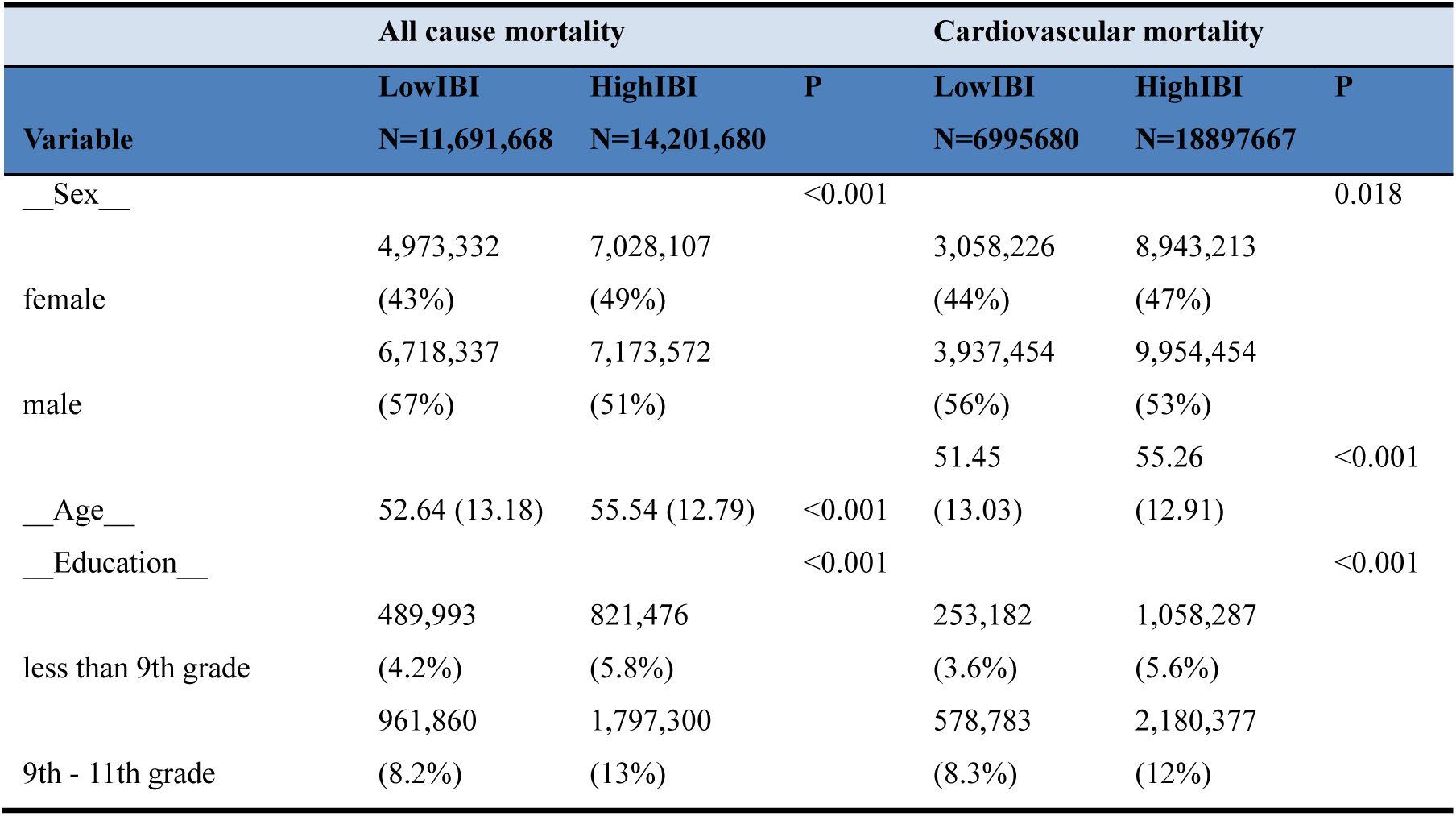
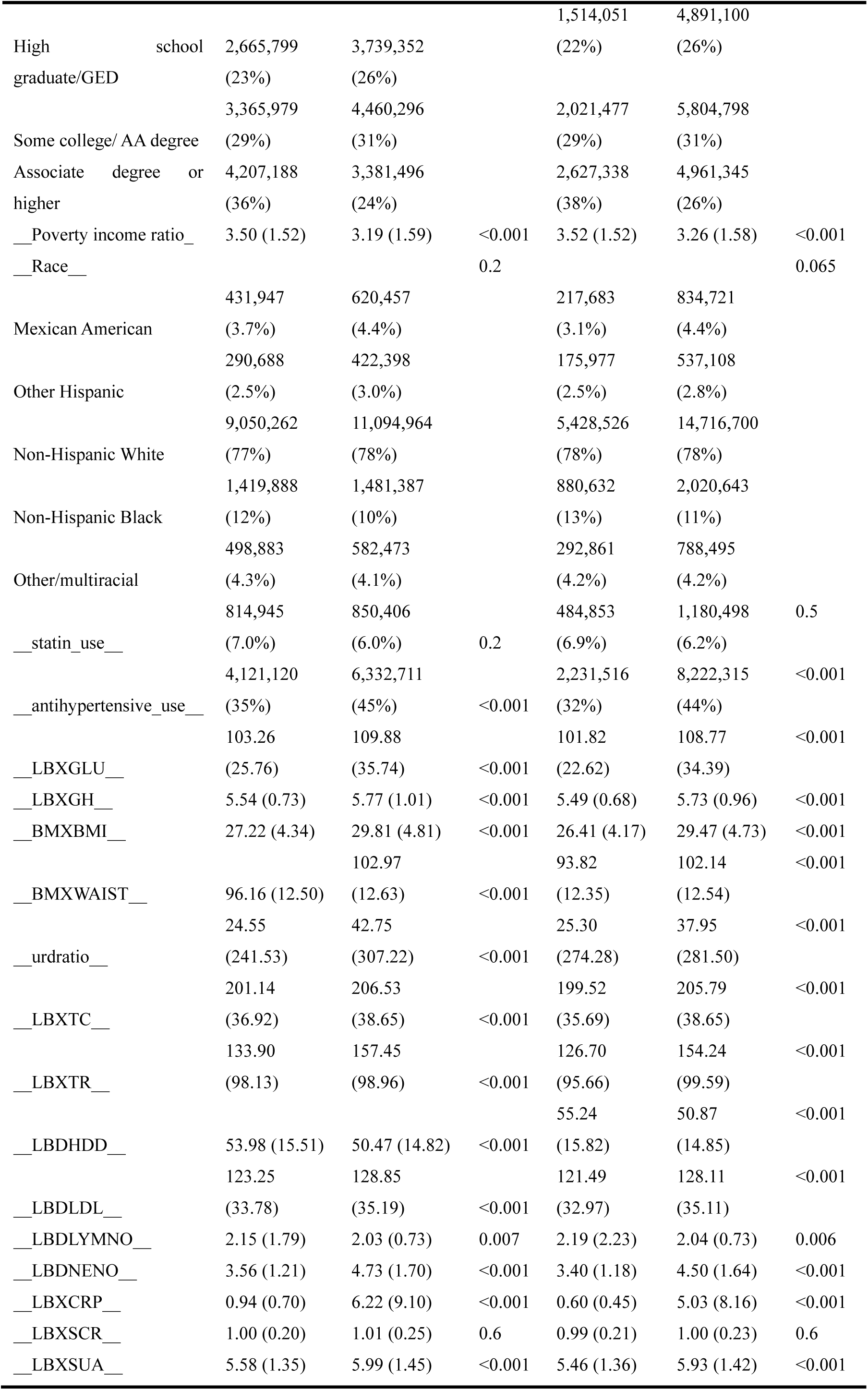
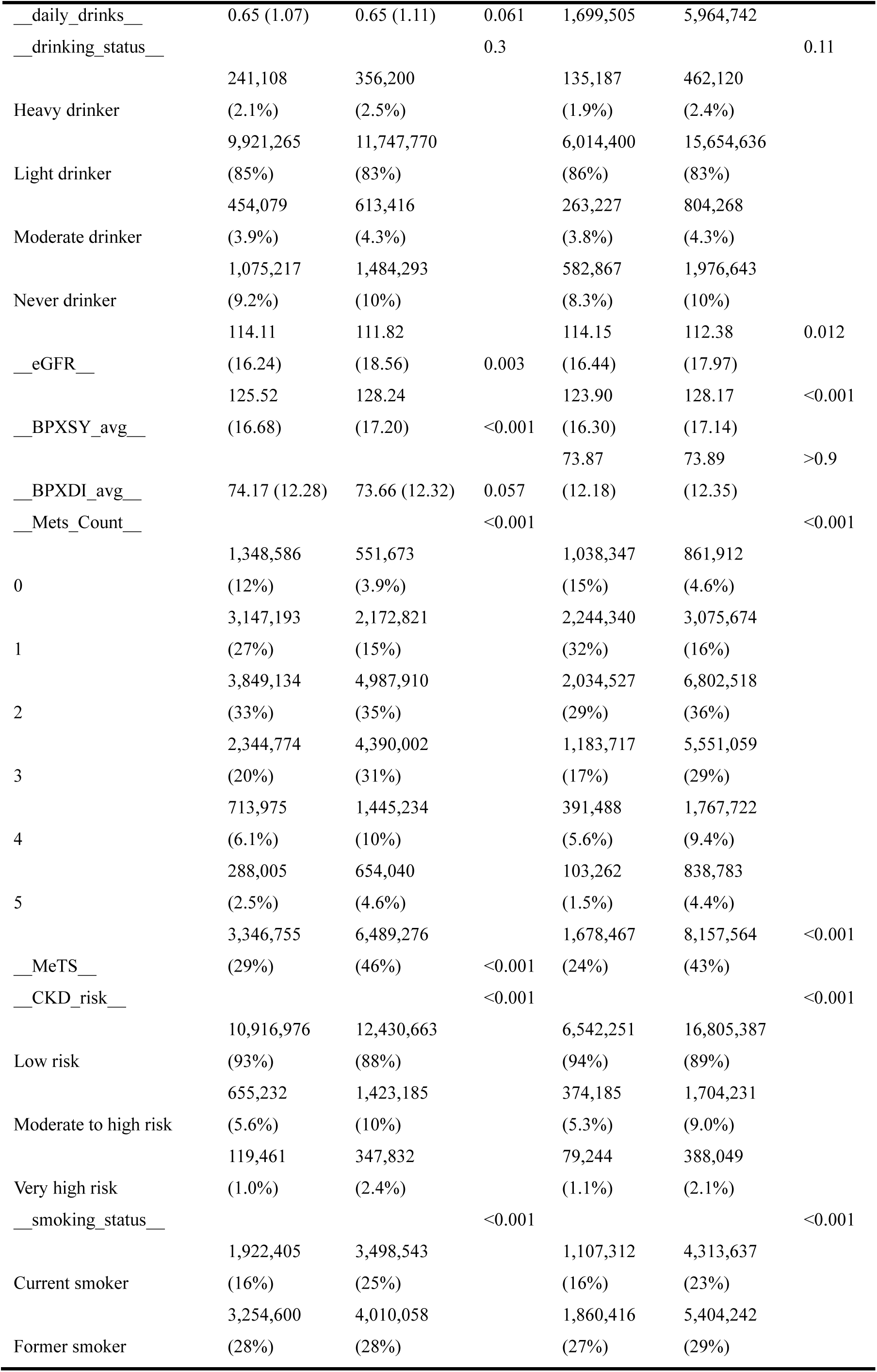
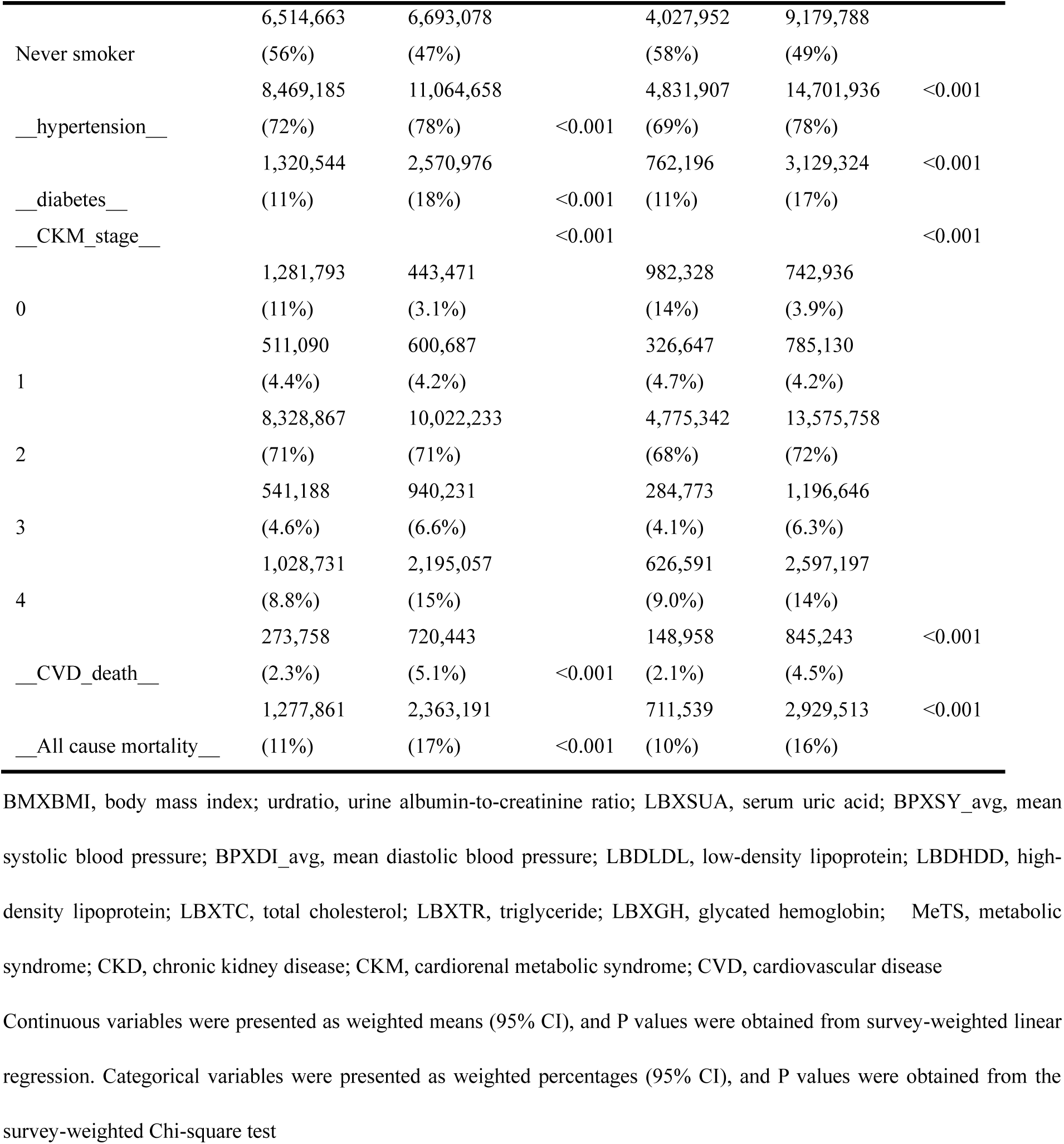
Baseline characteristics of participants with low and high ln_IBI, weighted for representativeness.

### Baseline Characteristics of Participants Categorized by Quartiles of ln_IBI

Table 2 demonstrates that when participants were categorized by quartiles of the log-transformed Inflammatory Burden Index (IBI), the Q4 group (highest IBI) exhibited a comprehensive health disadvantage compared to the Q1 group (lowest IBI). Specifically, Q4 had: (1) Lower socioeconomic status: Older age, lower educational attainment, and reduced income. (2) Significantly worse metabolic health: Poorer control of blood pressure, lipids, and blood glucose; more severe obesity; reduced renal function; and a higher prevalence meeting metabolic syndrome (MeTS) criteria. (3) Greater disease burden: Higher prevalence of hypertension, diabetes, MetS, elevated chronic kidney disease (CKD) risk, and advanced CKM stage, alongside a lower proportion in favorable health status. (4) Increased behavioral risk: Higher smoking prevalence. (5) Elevated mortality risk. These results further substantiate that a high IBI is a robust indicator of adverse health profiles and mortality outcomes in the CKM population.

**Table 2.**
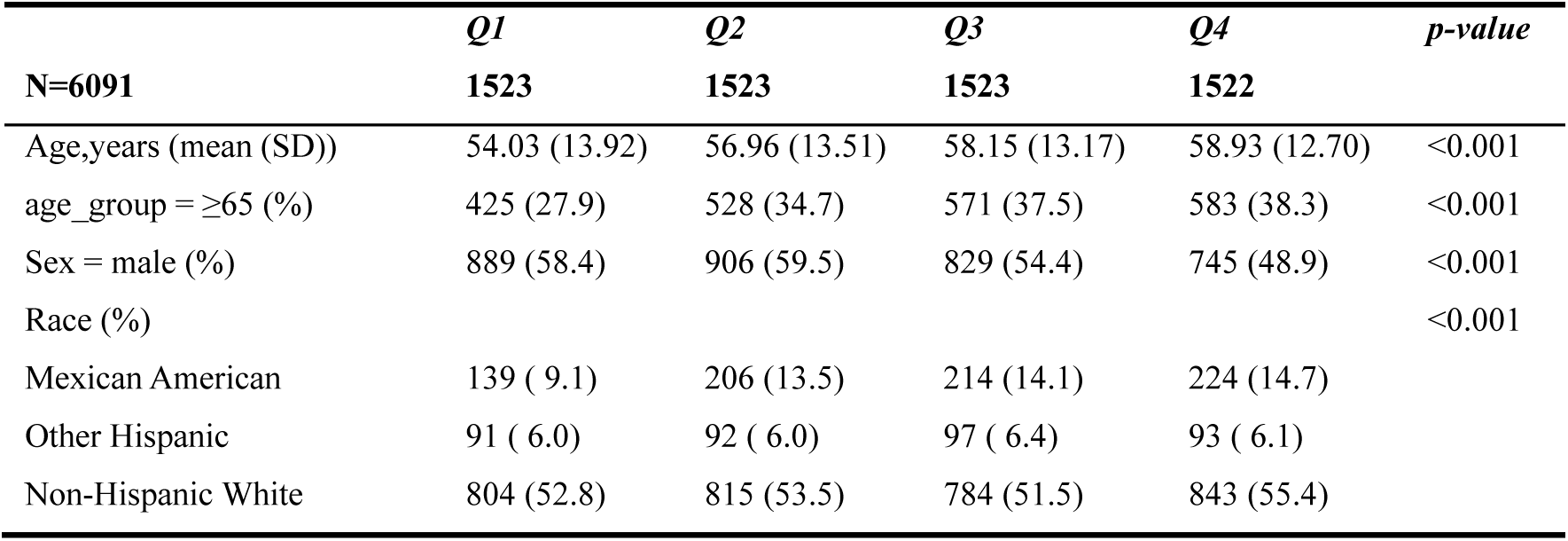
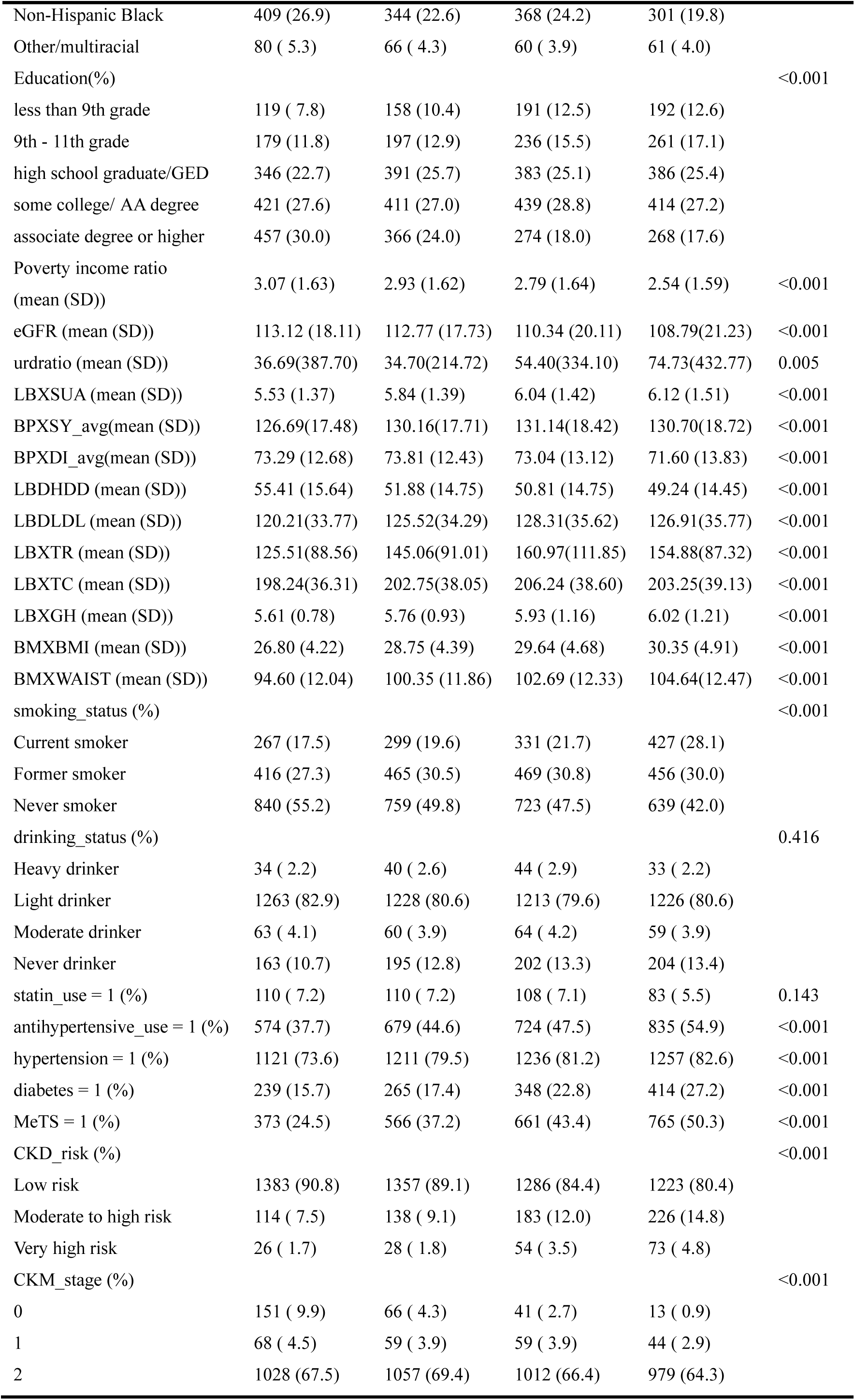
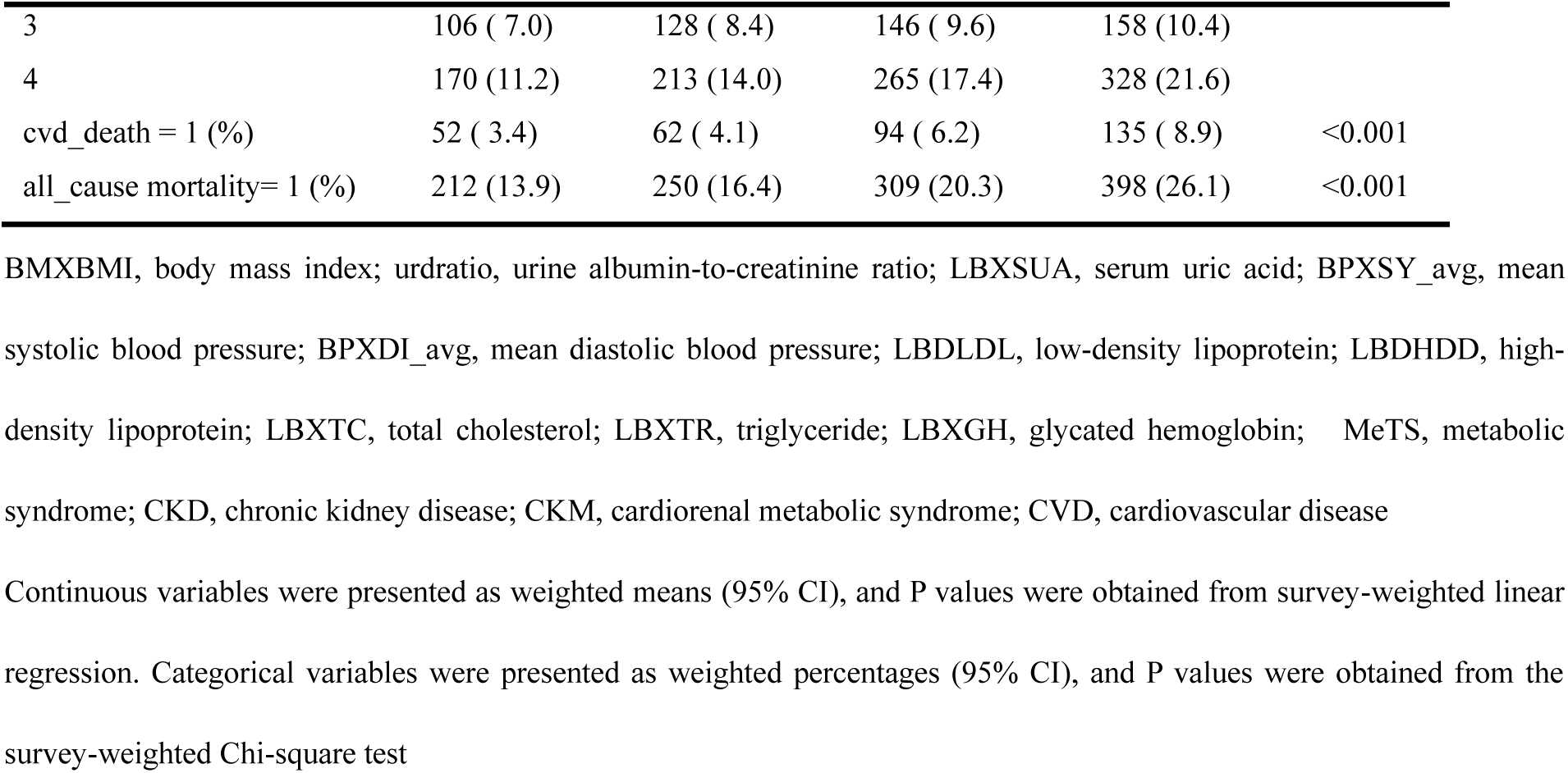
The baseline characteristics stratified by ln_IBI quartiles.

### Association of Inflammatory Burden index with all-cause mortality and cardiovascular mortality

Table 3 provides a summary of the relationships between the Inflammatory Burden Index (IBI) and both all-cause mortality and cardiovascular mortality. Prior to model development, an evaluation of multicollinearity among variables was conducted, and subsequently, three weighted Cox proportional hazards regression models were constructed. The natural logarithm of IBI (ln_IBI) was analyzed as a continuous variable in relation to all-cause and cardiovascular mortality. Weighted multivariable Cox regression analysis, which treated IBI as a continuous variable, demonstrated significant positive correlations between ln_IBI and both all-cause mortality and cardiovascular mortality. As shown in the table: In the unadjusted model (Model 1), each 1-unit increase in ln_IBI was associated with a significant 31% increased risk of all-cause mortality (HR = 1.31, 95% CI: 1.23–1.40, p < 0.001) and a 54% increased risk of cardiovascular mortality (HR = 1.54, 95% CI: 1.40–1.69, p < 0.001). After adjusting for age, sex, race/ethnicity, and PIR (Model 2), ln_IBI remained significantly positively associated with mortality risk (All-cause mortality: HR = 1.20, 95% CI: 1.12–1.28, p < 0.001; Cardiovascular mortality: HR = 1.44, 95% CI: 1.30–1.60, p < 0.001), although the effect sizes were attenuated compared to the unadjusted model. Following full adjustment for clinical and metabolic factors (Model 3), IBI remained an independent predictor of mortality risk (All-cause mortality: HR = 1.13, 95% CI: 1.05–1.21, p = 0.002; Cardiovascular mortality: HR = 1.35, 95% CI: 1.21–1.51, p < 0.001). These findings suggest that the relationship between the Inflammatory Burden Index (IBI) and cardiovascular mortality is more pronounced than its association with all-cause mortality among patients with CKM syndrome. Forest plots illustrate significant predictors from Model 3 for each outcome: For all-cause mortality (Figure 2), independent predictors included the Inflammatory Burden Index, older age, male sex, diabetes, high-risk CKD, and advanced CKM stage, while never smoking and higher income were protective factors. For cardiovascular mortality (Figure 3), strong predictors, besides the Inflammatory Burden Index, included male sex (HR = 2.10), diabetes (HR = 2.00), and elevated serum uric acid (HR = 1.19). The concordance index (C-index) progressively increased from Model 1 to Model 3 for both outcomes, indicating enhanced prediction accuracy and improved model discrimination with the inclusion of clinical variables. The proportional hazards assumption was satisfied for all three models, as confirmed by formal testing (detailed results in Supplemental Table 3 and Supplemental Table 4). Additionally, visual assessment using Schoenfeld residual plots for ln_IBI in Model 3 (Figure 4 and Supplemental Figure 5) demonstrated randomly distributed residuals, providing further evidence that the proportional hazards assumption was met.

**Figure 2.**
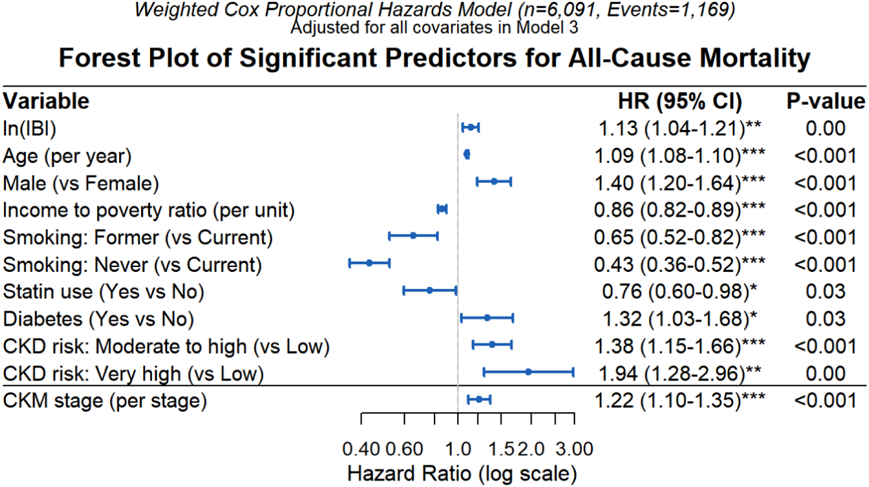
Significant effect variables in Model 3 of all - cause mortality. Data from n = 6091 participants in NHANES cohort. Hazard ratios (95% CI) derived from Cox proportional hazards regression using Wald tests for significance.

**Figure 3.**
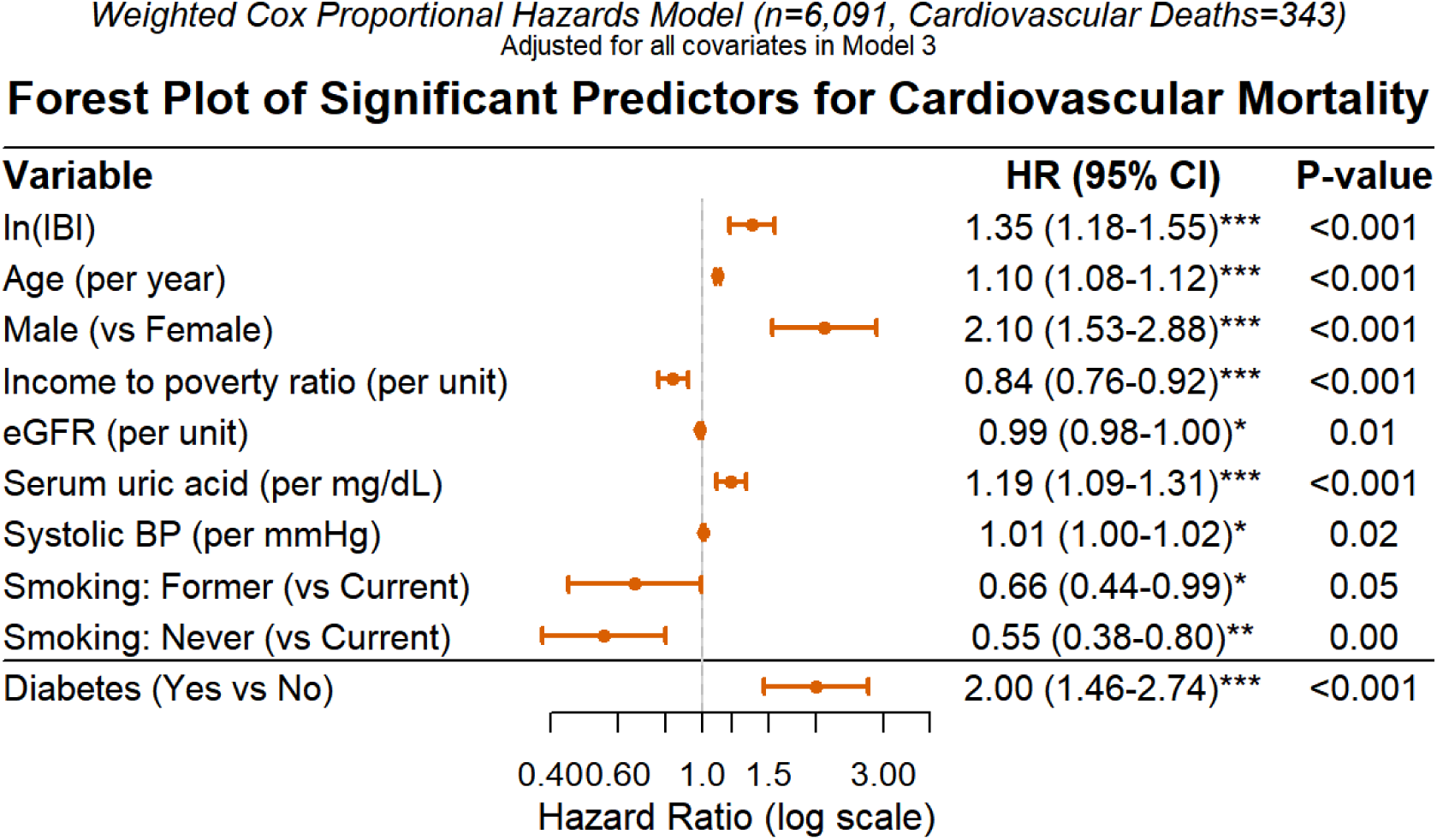
Significant effect variables in Model 3 of cardiovascular mortality. Data from n = 6091 participants in NHANES cohort. Hazard ratios (95% CI) derived from Cox proportional hazards regression using Wald tests for significance.

**Figure 4.**
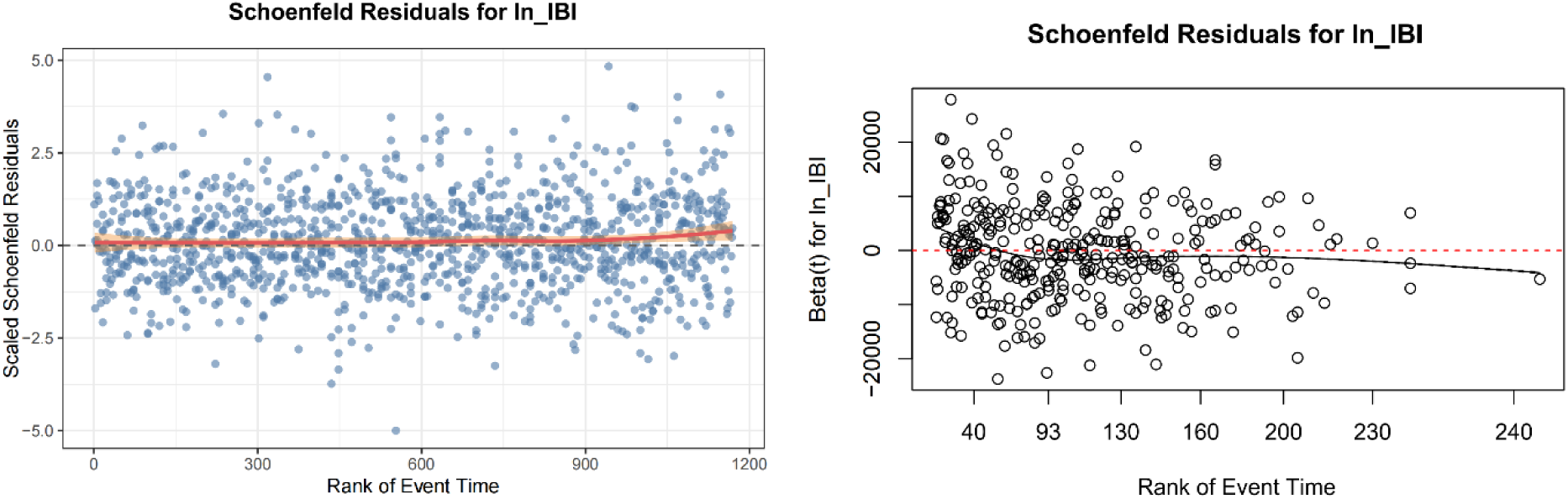
The left - hand figure shows the residual plot of ln_IBI in Model 3 of all - cause mortality, and the right - hand figure shows the residual plot of lb_IBI in Model 3 of cardiovascular mortality.

**Table 3.** Cox proportional hazards regression analysis of lnIBI concerning all-cause and cardiovascular mortality in a CKM population, weighted for representativeness.

| MODEL | All-cause mortality |  |  | Cardiovascular mortality |  |  |
| --- | --- | --- | --- | --- | --- | --- |
|  | HR_CI(95%) | P_VALUE | C-index | HR_CI(95%) | P_VALUE | C-index |
| Model1 | 1.31(1.23,1.40) | 4.5e-16 | 0.597 | 1.54(1.40,1.69) | 1.63e-16 | 0.662 |
| Model2 | 1.20(1.12,1.28) | 6.94e-08 | 0.815 | 1.44(1.30,1.60) | 3.63e-08 | 0.861 |
| Model3 | 1.13(1.05,1.21) | 1.79e-03 | 0.838 | 1.35(1.21,1.51) | 1.34e-05 | 0.88 |

Data from n = 6091 participants in NHANES cohort. Hazard ratios (95% CI) derived from Cox proportional hazards regression using Wald tests for significance.

We additionally assessed the predictive capability of the models through receiver operating characteristic (ROC) curves, with relevant results illustrated in Figures 5 and 6. Figure 5 displays the ROC curves for the weighted Cox regression models assessing the association between the Inflammatory Burden Index (IBI) and all-cause mortality at 3-year, 5-year, and 10-year intervals. The ROC curves demonstrate that Model 3 consistently exhibited the highest sensitivity and specificity across all thresholds. According to the Hosmer-Lemeshow criteria (where AUC > 0.7 indicates acceptable discrimination and AUC > 0.8 indicates excellent discrimination), Model 3 demonstrated excellent discriminatory ability. Figure 6 illustrates the change in the area under the curve (AUC) values over time for the three models predicting cardiovascular disease (CVD) mortality. It is evident that Model 3 had the best predictive performance, maintaining the highest AUC values, consistently greater than 0.8. This indicates that the comprehensive model incorporating IBI, demographic factors, and clinical indicators possesses the optimal predictive power for mortality risk.

**Figure 5.**
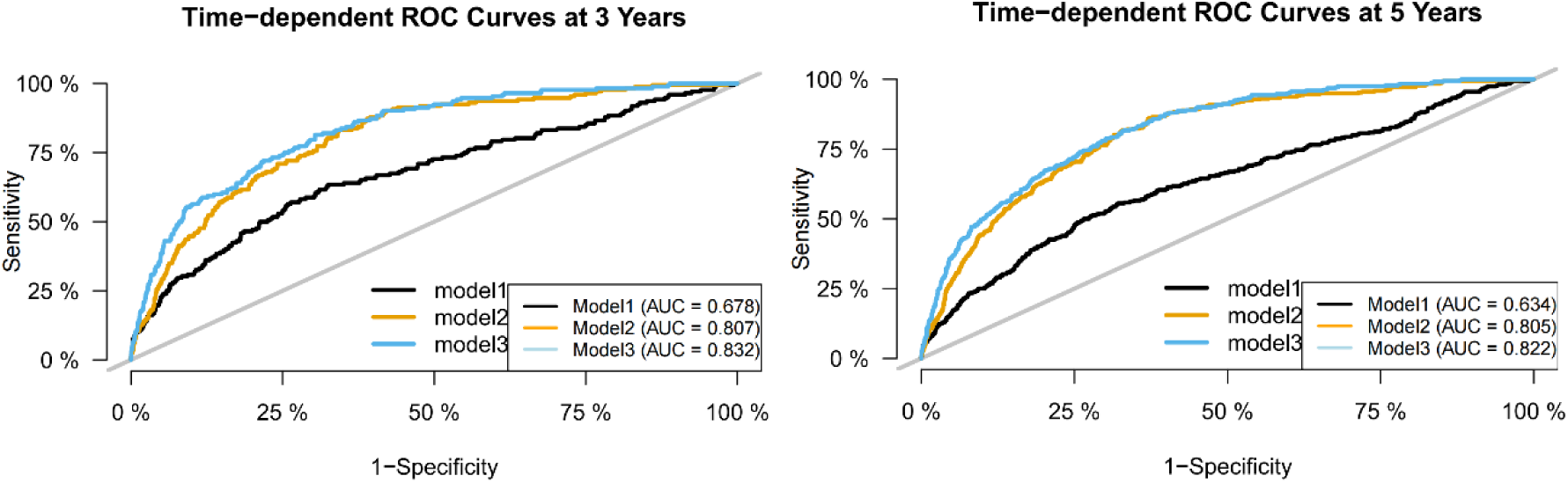
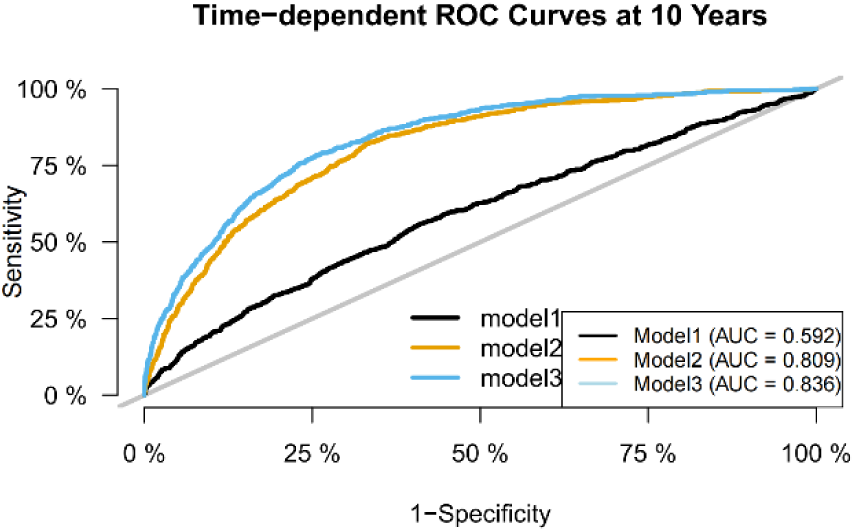
ROC curve of the weighted Cox regression model between IBI and all-cause mortality.

**Figure 6.**
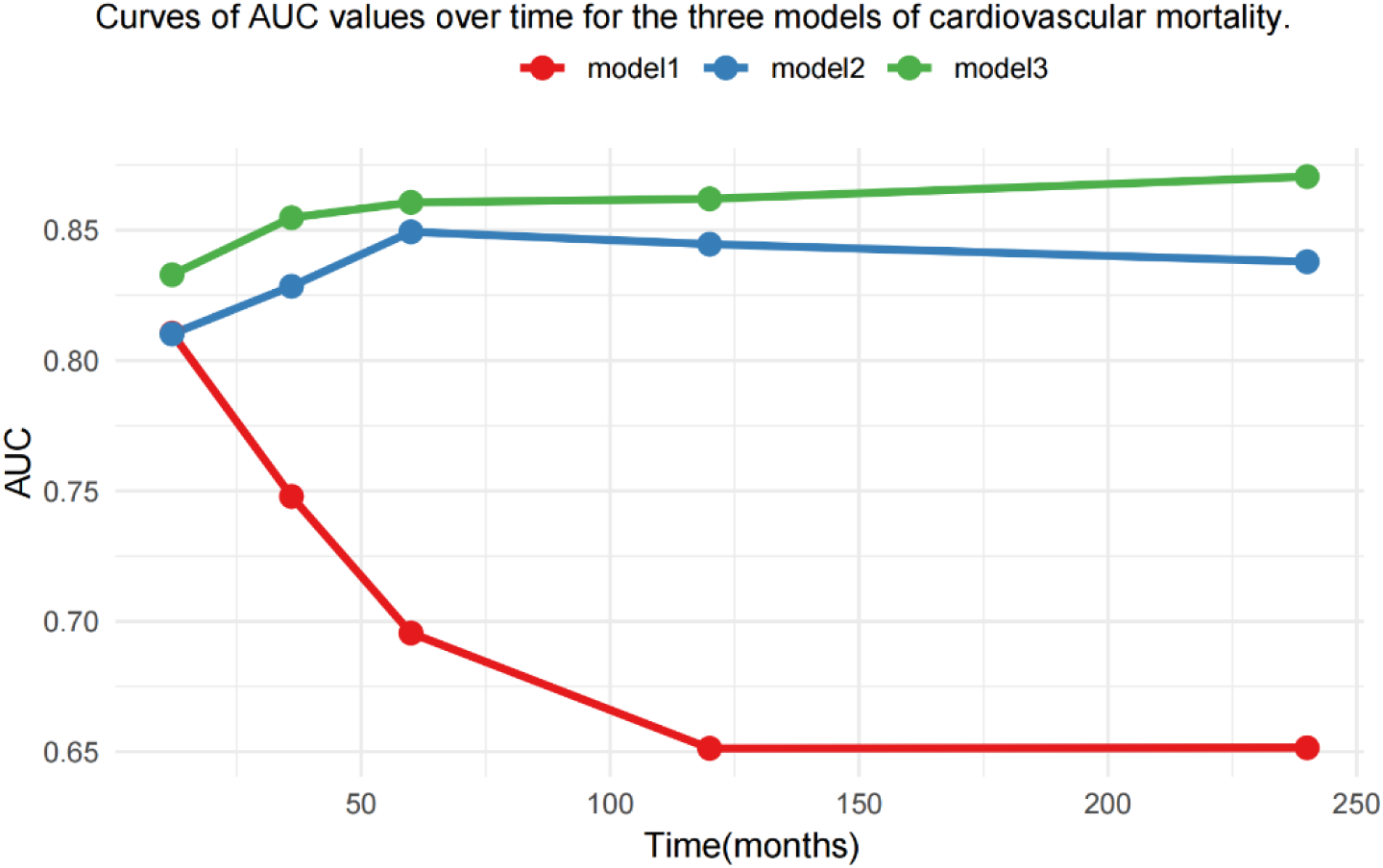
Curves of AUC values over time for the three models of cardiovascular mortality.

### Nonlinear relationship of inflammatory burden index with all-cause mortality and cardiovascular mortality, weighted for representativeness

Due to the right-skewed distribution of the Inflammatory Burden Index (IBI) data, we performed weighted Cox regression analyses using quartiles of the log-transformed IBI (ln_IBI) to assess associations with all-cause and cardiovascular mortality in the CKM population (Table 4). Model 1 (Unadjusted): Elevated IBI levels (Q3, Q4) showed strong, significant associations with increased risks of both all-cause and cardiovascular mortality compared to the reference quartile (Q1). A clear dose-response relationship was evident, with Hazard Ratios (HRs) progressively increasing across higher quartiles. The Q4 group exhibited the highest risk. Model 2 (Demographically Adjusted): The strength of the association between IBI and mortality risk was significantly attenuated. Associations for Q2 and Q3 groups were largely eliminated (HRs close to 1, non-significant p-values). Only the Q4 group retained a significant positive association with both all-cause and cardiovascular mortality, although HR values were substantially reduced compared to Model 1. Model 3 (Fully Adjusted): After adjusting for comprehensive clinical biomarkers and potential mediating variables, the association between IBI and all-cause mortality became non-significant (Q4 HR = 1.14, 95% CI: 0.93–1.43, p = 0.294). Notably, even in this maximally adjusted model (Model 3), individuals in the highest IBI quartile (Q4) had a significantly higher risk of cardiovascular mortality compared to those in the lowest quartile (Q1) (HR = 1.77, 95% CI: 1.18–2.67, p = 0.008). This analysis demonstrates that the highest level of IBI (Q4) is a robust predictor of cardiovascular death. Even after maximal adjustment for a wide range of potential confounders and clinical indicators (Model 3), the cardiovascular mortality risk in the Q4 group remained significantly elevated by 77% (HR = 1.77, 95% CI: 1.18–2.67) compared to the Q1 group. In contrast, associations for lower IBI levels (Q2, Q3) with cardiovascular mortality became non-significant after adjustment. For all-cause mortality, the initially observed significant risk increases associated with elevated IBI (Q3, Q4) became non-significant after adjusting for lifestyle and underlying diseases (Model 2), and particularly after incorporating detailed clinical biomarkers and medication history (Model 3). However, our previous analysis treating IBI as a continuous variable revealed a significant positive association between ln_IBI and all-cause mortality in Model 3. This apparent contradiction may arise from a significant non-linear relationship between IBI and all-cause mortality in the CKM patients. Thus, we conducted a further analysis of the association between IBI and mortality by means of restricted cubic splines (RCS).

**Table 4.**
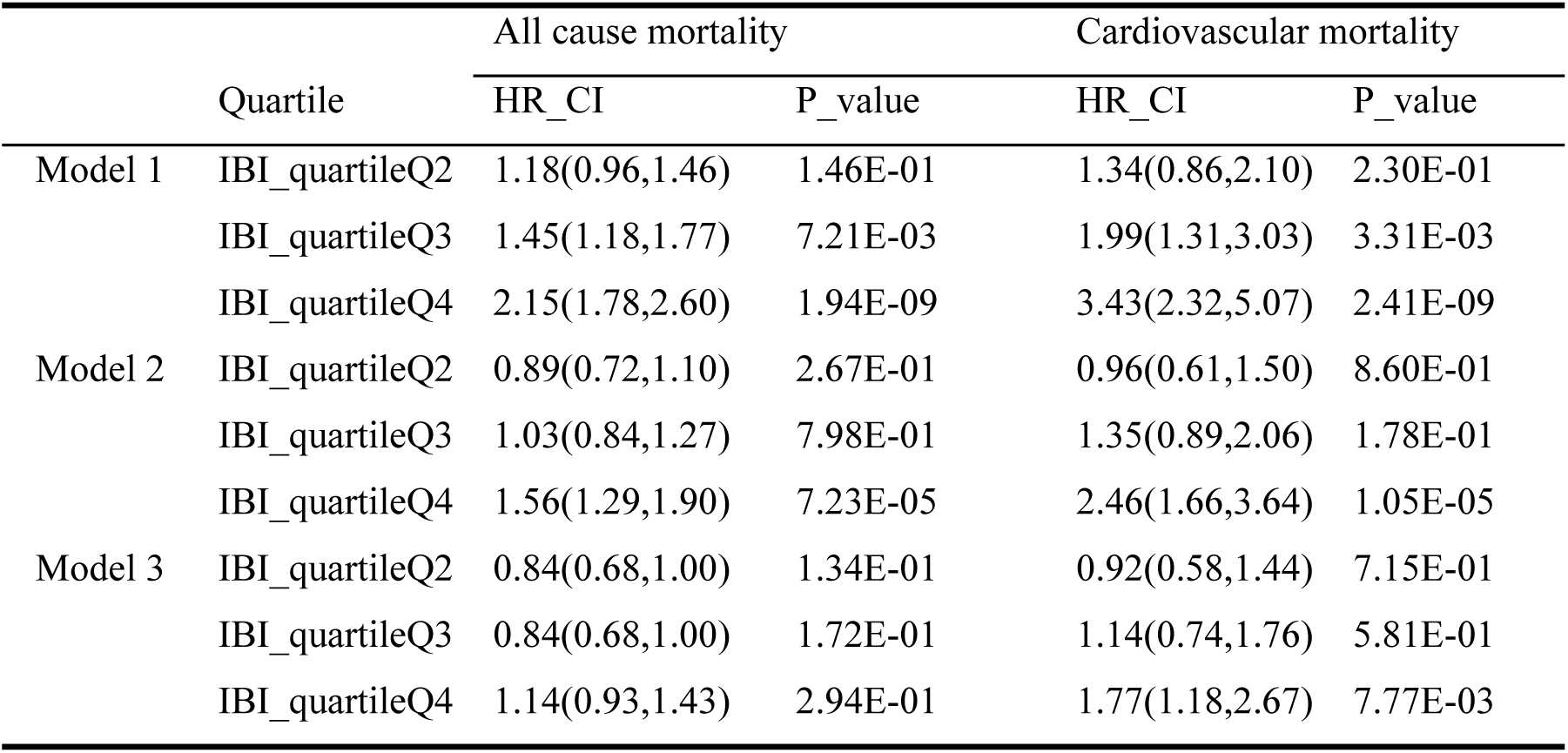
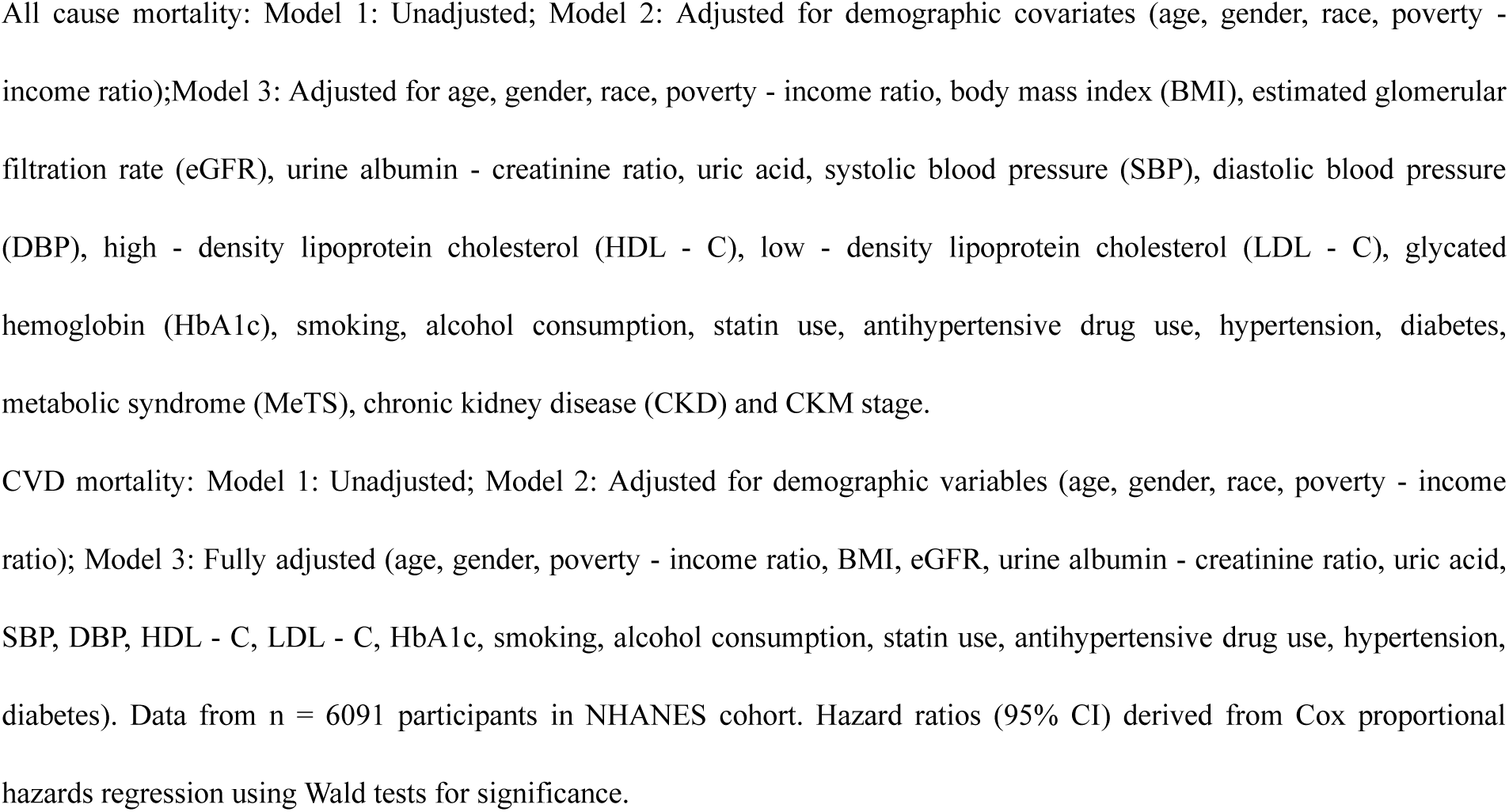
Weighted Cox regression analysis of quartiles of ln_IBI and mortality in the CKM population.

Figure 7 depicts the dose-response curve for IBI and all-cause mortality. The curve exhibited a “U-shaped” relationship, indicating complex non-linearity. The curve reached a nadir (point of lowest risk) at IBI = 3.357 (HR = 0.479, 95% CI: 0.251–0.914). HR values increased gradually on both sides of this nadir, with a steeper slope on the right side, suggesting a greater long-term hazard associated with higher IBI. The curve intersected the HR=1 line at two points (IBI = 0.0864 and IBI = 52.49), defining a relatively safe range (IBI 0.0864–52.49) where mortality risk was ≤ baseline level (HR ≤ 1). Figure 8 shows the dose-response curve for IBI and cardiovascular mortality. An inflection point was identified at IBI = 1.684. Left of this inflection point, the curve was relatively flat, indicating minimal change in risk. Right of the inflection point, the risk increased sharply. The RCS analysis suggests a clear threshold effect: IBI significantly increased cardiovascular mortality risk only when exceeding 1.684 (HR at inflection = 0.764, 95% CI: 0.26–2.24), consistent with the multi-model analysis results above. (Detailed RCS analysis results are provided in Supplemental Tables 5 and 6). The “J-shaped” or “U-shaped” characteristic explains the apparent contradiction in the earlier all-cause mortality analyses: a significant positive association for ln_IBI as a continuous variable, yet non-significant associations for the quartiles (especially Q4) in Model 3. The RCS analysis revealed that the IBI-associated excess risk for all-cause mortality is predominantly concentrated at the very highest end of the distribution. When IBI is discretized into quartiles, the highest quartile (Q4) likely contains individuals with significantly elevated risk (concentrated in the upper segment of Q4) alongside individuals whose risk is relatively closer to the reference level (lower segment of Q4). This dilution of the average risk effect within Q4 makes it harder to achieve statistical significance for the quartile variable. Conversely, continuous variable analysis preserves the full information of IBI and is more sensitive for detecting non-linear associations, particularly effects at the distribution extremes. Categorical analysis loses information and reduces statistical power to detect risk clustering within Q4, especially among individuals with the most extreme values.

**Figure 7.**
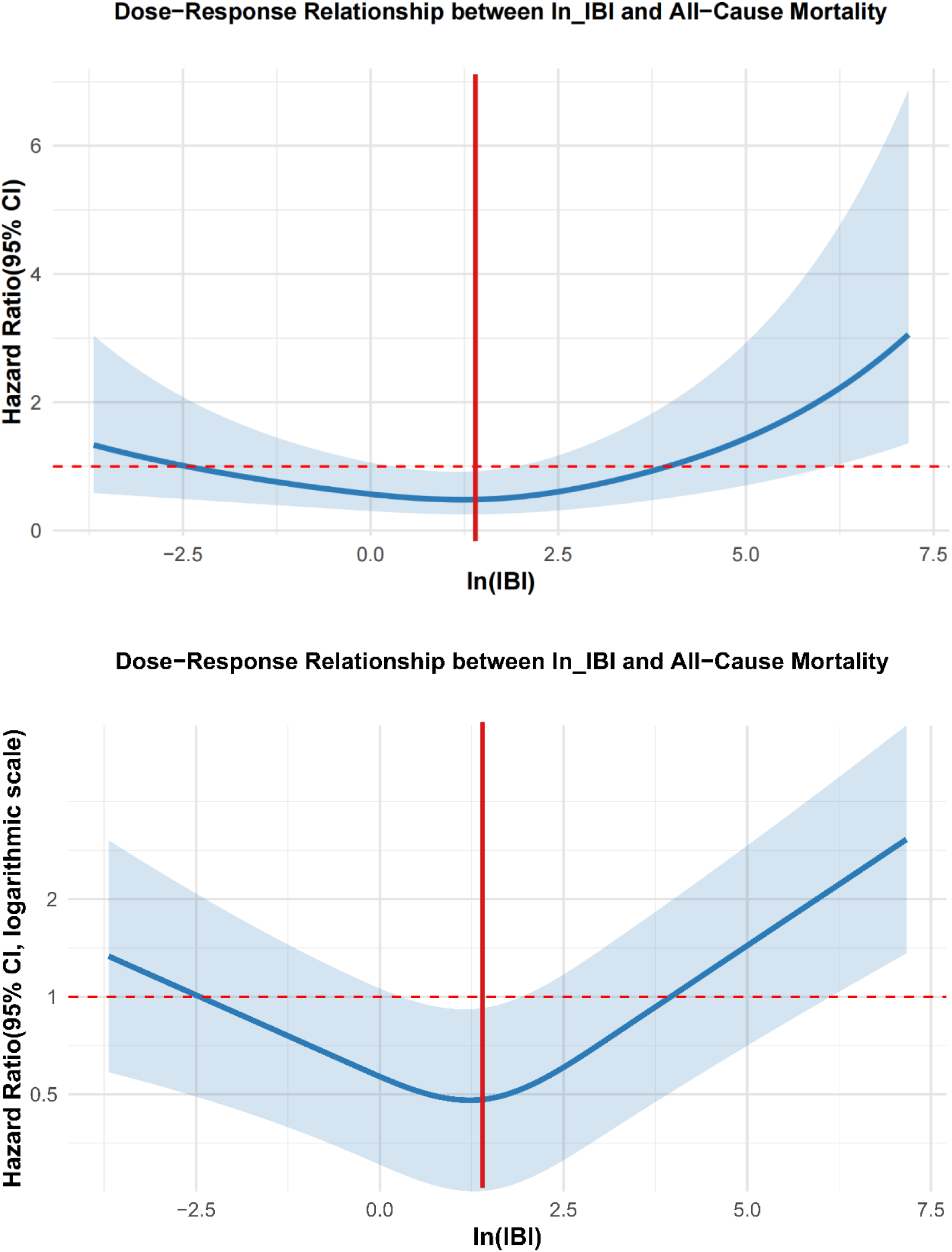
Dose−Response Relationship between IBI and All cause Mortality. Restricted cubic spline curve with 3 knots (AIC-optimized). The curve reached a nadir (point of lowest risk) at IBI = 3.357 (HR = 0.479, 95% CI: 0.251–0.914).The curve intersected the HR=1 line at two points (IBI = 0.0864 and IBI = 52.49), defining a relatively safe range (IBI 0.0864–52.49) where mortality risk was ≤ baseline level (HR ≤ 1). Nonlinearity: Likelihood ratio test (P = 0). Data from n = 6091 participants in NHANES cohort (All-cause deaths=1169).

**Figure 8.**
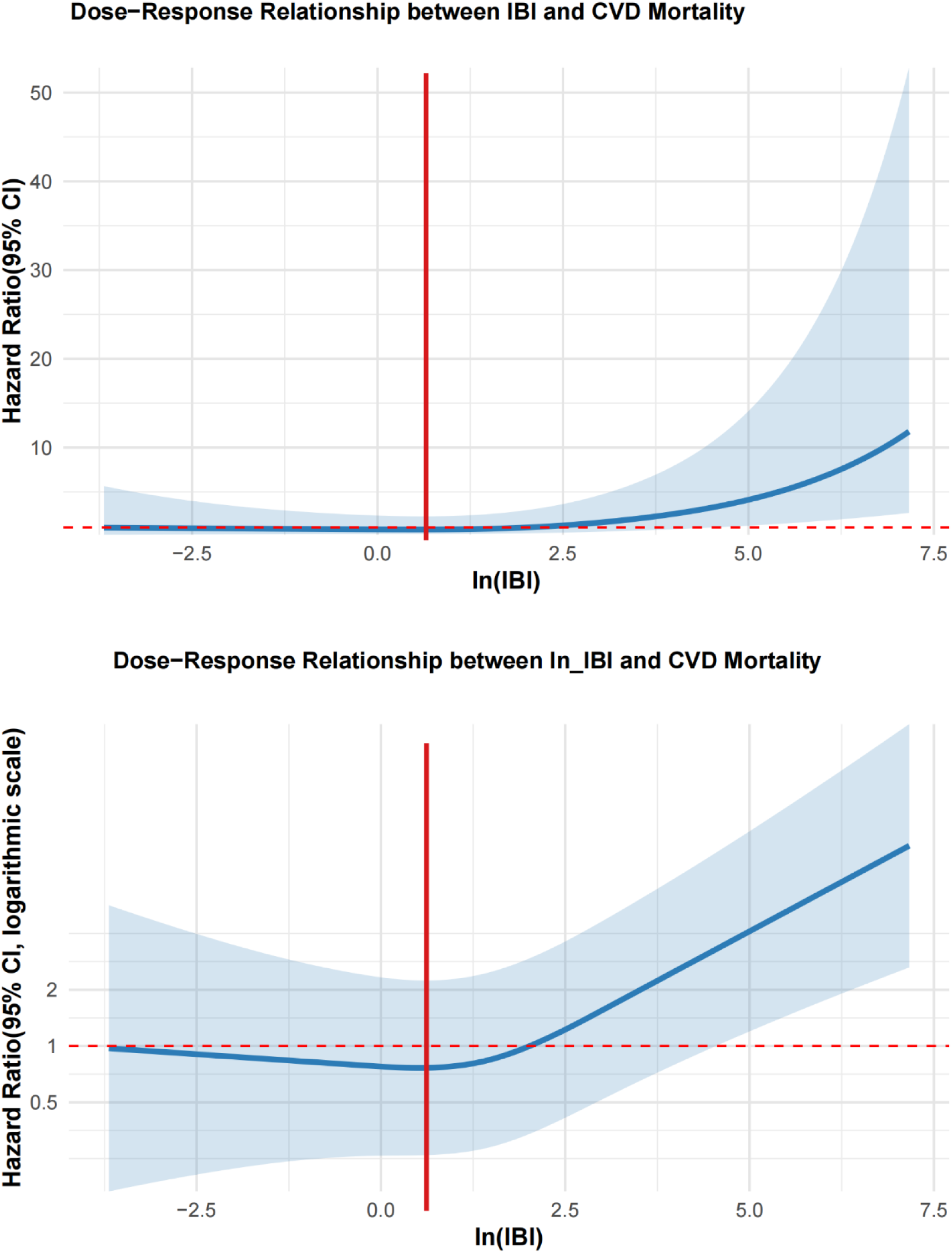
Dose−Response Relationship between IBI and CVD Mortality. Restricted cubic spline curve with 3 knots (AIC- optimized). An inflection point was identified at IBI = 1.684(HR at inflection = 0.764, 95% CI: 0.26–2.24). Nonlinearity: Likelihood ratio test (P = 0.012). Data from n = 6091 participants in NHANES cohort (CVD deaths = 343).

### Subgroup analysis, stratified analysis, and sensitivity analysis

Figure 9 presents a forest plot of subgroup analysis results for the association between the Inflammatory Burden Index (IBI) and all-cause mortality in the CKM population. Subgroup analyses were stratified according to age, sex, racial/ethnic background, smoking status, alcohol consumption habits, disease status (including hypertension, diabetes, and metabolic syndrome), chronic kidney disease (CKD) risk stage, and CKM stage, with adjustments made for relevant covariates. (1) Age: The association between IBI and all-cause mortality was significantly stronger among individuals aged ≥65 years (HR = 1.15, 95% CI: 1.09–1.22, p < 0.001), suggesting a more pronounced impact of IBI on mortality risk in older adults, potentially due to reduced physiological reserve. In contrast, no statistically significant association was observed in those aged <65 years (HR = 1.04, 95% CI: 0.95–1.14, p = 0.350), indicating that age may act as an effect modifier. (2) Sex: Both males (HR = 1.12, 95% CI: 1.05–1.19, p < 0.001) and females (HR = 1.11, 95% CI: 1.03–1.21, p = 0.010) exhibited increased mortality risk with higher IBI. (3) Race: Significant racial/ethnic disparities were evident. Associations were stronger in Mexican Americans (HR = 1.27, 95% CI: 1.09–1.48, p = 0.002) and Non-Hispanic Blacks (HR = 1.13, 95% CI: 1.06–1.21, p < 0.001). However, no significant association was found in other subgroups like Other Hispanics (HR = 1.24, 95% CI: 0.94–1.63, p = 0.130), potentially reflecting race-specific health disparities (e.g., distribution of underlying diseases, healthcare access). (4) Smoking Status: A positive association between IBI and the risk of mortality was identified in current smokers, former smokers, and individuals who have never smoked. Notably, current smokers exhibited the largest effect size, suggesting that smoking may synergistically amplify the mortality risk associated with elevated IBI. (5) Disease Status: Hypertension: The association was more pronounced in individuals with hypertension (HR = 1.11, 95% CI: 1.05–1.17, p < 0.001), indicating that hypertension is another synergistic factor exacerbating IBI-associated mortality risk. MetS and CKD: Stronger associations were observed in individuals with metabolic syndrome (MetS) (HR = 1.15, 95% CI: 1.06–1.25, *p* < 0.001) and those at very-high-risk CKD (HR = 1.35, 95% CI: 1.07–1.71, *p* = 0.010). The highest risk was seen in CKD stage 4 individuals (HR = 1.18, 95% CI: 1.08–1.29, *p* < 0.001), indicating that metabolic dysregulation and impaired renal function amplify the detrimental impact of IBI on survival. Overall, the association between IBI and all-cause mortality exhibited significant population heterogeneity, with elevated risks observed in older adults, specific racial/ethnic groups, smokers/moderate drinkers, and individuals with comorbidities like hypertension, MeTS, or advanced CKD.

**Figure 9.**
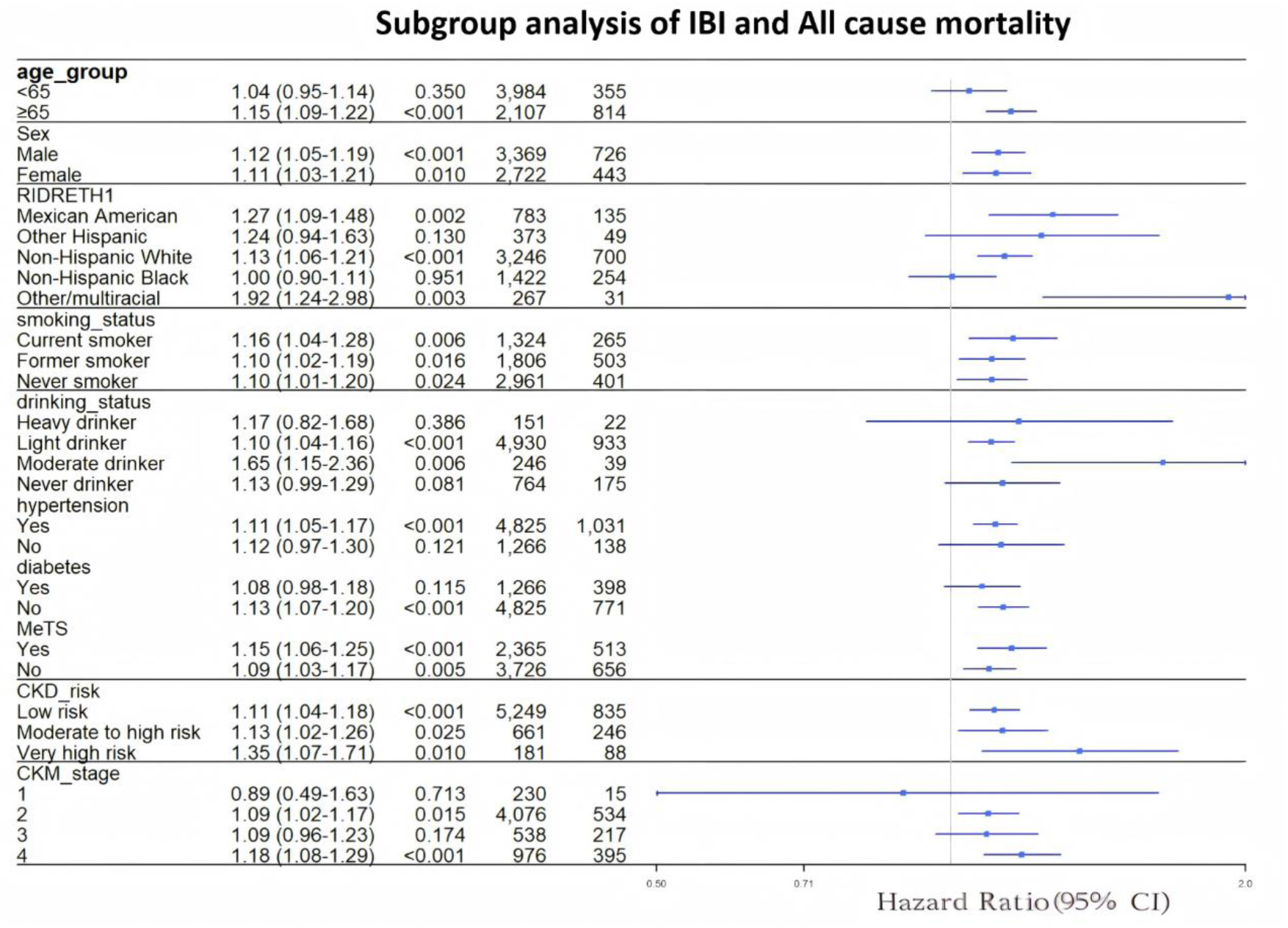
Subgroup analysis of the association between IBI and all-cause mortality in patients with CKM syndrome. Statistical tests: Within-subgroup effects: Wald tests for ln(IBI) coefficient. Interaction effects: Wald-type F-tests with design-based variance estimation.

Figure 10 presents the forest plot for subgroup analysis of IBI and cardiovascular mortality. It demonstrates that the association between IBI and CVD mortality also exhibits population heterogeneity, influenced by sex, race, smoking status, and comorbidities. Specifically, female sex, current smoking, diabetes, and metabolic abnormalities were identified as factors that synergistically amplified the cardiovascular mortality risk associated with elevated IBI in those with CKM syndrome. Figure 11 presents the forest plot for stratified analysis of IBI and all-cause mortality. It shows that older age, male sex, smoking, and Non-Hispanic Black race augmented the mortality risk effect of IBI. Comorbidities such as hypertension, diabetes, and high-risk CKM syndrome further strengthened the association between IBI and mortality risk. Stratified analysis for IBI and cardiovascular mortality was not performed due to insufficient event numbers within strata.

**Figure 10.**
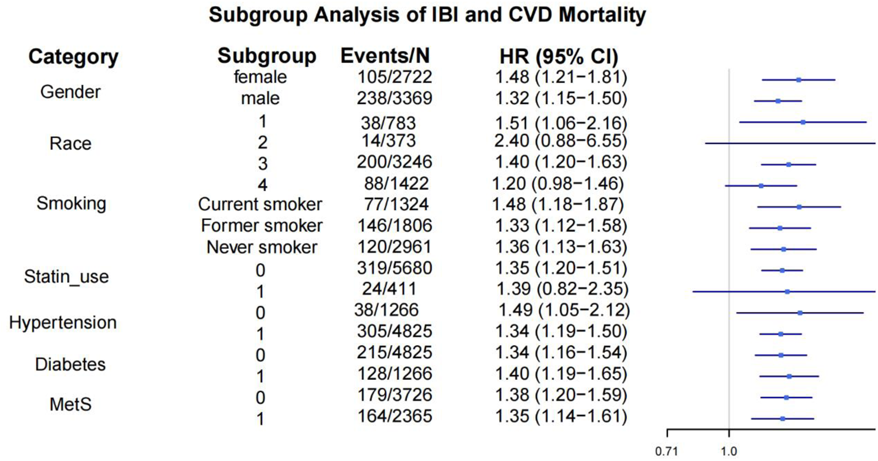
Subgroup analysis of the association between IBI and cardiovascular mortality in patients with CKM syndrome. Within-subgroup effects: Wald tests for ln(IBI) coefficient. Interaction effects: Wald-type F-tests with design-based variance estimation.

**Figure 11.**
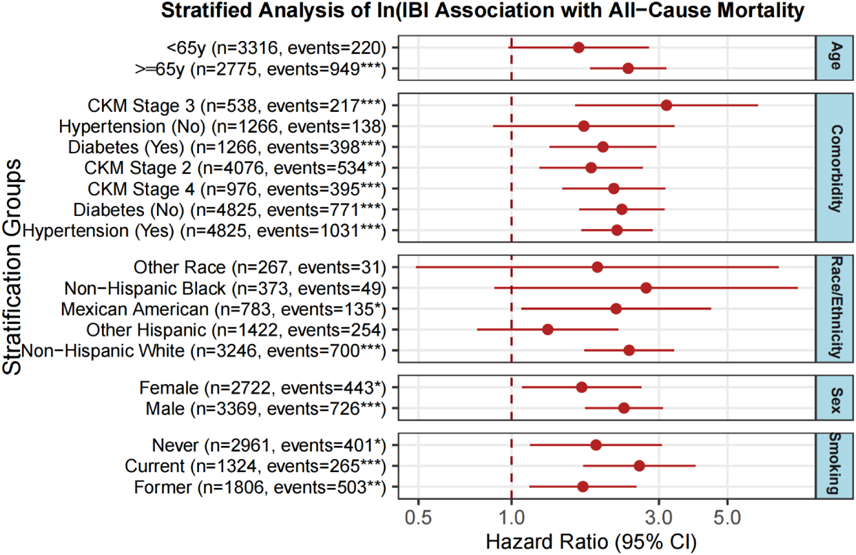
Stratified analysis of the association between IBI and all-cause mortality in patients with CKM syndrome. Hazard ratios (95% CI) derived from Cox proportional hazards regression using Wald tests for significance.

Additionally, sensitivity analyses for the association between IBI and all-cause mortality are presented in Table 5. Both sets of sensitivity analyses consistently demonstrated a significant positive association between IBI and all-cause mortality (HR > 1 and p < 0.05). While the effect size slightly attenuated after excluding participants with pre-existing CVD (HR decreased from 1.115 to 1.108), the association remained statistically significant. This confirms that IBI is a robust risk factor for all-cause mortality, and its detrimental effect persists significantly even after excluding individuals with established CVD.

**Table 5.** Sensitivity analysis of IBI and All cause mortality.

| Analysis | HR | Lower_CI | Upper_CI | P_value |
| --- | --- | --- | --- | --- |
| Exclude CVD | 1.107586 | 1.001517 | 1.224889 | 0.046648305 |
| Complete Cases | 1.115254 | 1.034513 | 1.202296 | 0.004442739 |

The group with CVD deaths excluded had n = 5748, and the group with complete cases had n = 6091. Hazard ratios (95% CI) derived from Cox proportional hazards regression using Wald tests for significance.

## 4 Discussion

Our study is the first to demonstrate that the novel inflammatory biomarker, Inflammatory Burden Index (IBI), has an independent and significant association with both all-cause mortality and cardiovascular mortality in the population with CKM syndrome. Of particular importance, this association exhibits complex non-linear characteristics and is significantly influenced by demographic features and comorbidity status. These findings provide new epidemiological evidence for understanding the core role of inflammation in the progression of CKM syndrome.

### 4.1 Summary of Key Findings

This study is the first to systematically evaluate the prognostic value of the composite inflammatory marker Inflammation-Based Index (IBI) in a large, nationally representative population with Cardiometabolic-Kidney (CKM) syndrome. The core findings are as follows:

4.1.1 To explore the association between the novel biomarker, the Inflammatory Burden Index (IBI), and mortality in the CKM syndrome population, we initially conducted weighted Cox regression analyses, considering IBI as a continuous variable. These analyses revealed significant positive associations between IBI and both all-cause and cardiovascular mortality, establishing IBI as an independent risk factor for these outcomes in the CKM population. Subsequently, ROC curve analysis showed that IBI remained a significant predictor of increased all-cause and cardiovascular mortality risk in the CKM population, even after comprehensive adjustment for age, sex, race/ethnicity, poverty-income ratio, clinical biomarkers (fasting glucose, LDL-C, HDL-C, HbA1c, UACR, etc.), disease status (hypertension, diabetes, metabolic syndrome), and medication use. Further analysis using quartiles of the log-transformed IBI (ln_IBI) revealed a significant non-linear relationship between IBI and mortality risk in the CKM population. Restricted cubic spline (RCS) analysis confirmed this non-linearity: The RCS curve for IBI and all-cause mortality exhibited a nadir (point of lowest risk) and an optimal low-risk range. The RCS curve for IBI and cardiovascular mortality also displayed a nadir. To assess potential synergistic effects of covariates and mitigate confounding, we conducted subgroup, stratified, and sensitivity analyses.

4.1.2 The association with IBI exhibited significant population heterogeneity. Elevated risks were particularly pronounced in older adults, specific racial/ethnic groups, smokers/moderate drinkers, and individuals with comorbidities like hypertension, MetS, or advanced CKD. Association heterogeneity was also observed, influenced by sex, race, smoking status, and comorbidities. Female sex, current smoking, diabetes, and metabolic abnormalities were identified as factors that synergistically amplified the cardiovascular mortality risk associated with elevated IBI. The significant positive association between IBI and all-cause mortality persisted even after excluding participants with pre-existing CVD, confirming IBI as a robust risk factor.

### 4.2 Comparison and Integration with Existing Evidence

The IBI, a novel biomarker initially proposed based on observations of elevated pro-inflammatory cytokines (e.g., C-reactive protein, CRP) and inflammatory cells (e.g., neutrophils, lymphocytes) in synovial tissue, synovial fluid, and peripheral blood of osteoarthritis patients^18–20^, quantifies systemic inflammation. Cardiovascular-Kidney-Metabolic (CKM) syndrome, defined as the pathophysiological interplay among the cardiovascular system, kidneys, and metabolic risk factors leading to multi-organ dysfunction and complex clinical manifestations^2^, is fundamentally rooted in metabolic dysregulation (central obesity, hypertension, insulin resistance, dyslipidemia)^29^, a process intrinsically linked to inflammation^30^. CRP is an independent predictor of metabolic syndrome and cardiovascular events. A review concluded that CRP demonstrates a dose-response relationship with cardiovascular risk, can identify subgroups among metabolic syndrome patients who are at heightened risk, and likely functions as a biomarker for atherosclerotic vascular inflammation^31^. A prospective cohort study revealed that CRP is independently linked to an elevated long-term risk in patients with heart failure^32^. Another prospective cohort study conducted in CVD patients at the University Medical Center Utrecht, Netherlands, found that higher CRP, as an indicator of chronic low-grade inflammation, was independently associated with increased risk of CVD recurrence and mortality, unaffected by traditional CVD risk factors and preventive medication use; this association persisted for 15 years after the initial CRP measurement^33^. CRP is also a risk factor and predictive biomarker for diabetes A 7-year follow-up study of adults in Tromsø, Norway, showed that CRP is associated with future development of diabetes, can be used for diabetes risk stratification, and this association is consistent across sex, BMI, hypertension, and abdominal obesity^34^. In patients with stage 3 and 4 chronic kidney disease, CRP is a risk factor for CKD and an independent risk factor for all-cause mortality^35^. Elevated serum CRP levels are likely to facilitate the infiltration of inflammatory cells (such as neutrophils and macrophages) and stimulate the secretion of cytokines, chemokines, and transforming growth factor-β1 (TGF-β1) from impaired kidneys, thereby contributing to the progression of renal inflammation and fibrosis^36^. Research has shown that the neutrophil-to-lymphocyte ratio (NLR) can function as an independent prognostic indicator for all-cause mortality in patients with coronary heart disease complicated by hypertension^37^.

Aligned with this large amount of previous work, our weighted Cox regression analyses confirm that the Inflammatory Burden Index (IBI) is a significant risk factor for both all-cause mortality and cardiovascular mortality in the CKM syndrome population. This association is particularly pronounced among high-risk subgroups, including the elderly, individuals with obesity, those with multiple comorbidities (hypertension, diabetes, metabolic syndrome), and smokers. A key advancement of this study lies in the fact that IBI, as a composite marker integrating innate immunity (neutrophils), adaptive immunity (lymphocytes), and CRP, may more comprehensively capture the complex inflammatory network status in CKM syndrome. Compared with individual markers, IBI provides stronger prognostic discriminatory power, which is evidenced by the increase in the model’s C-index following the inclusion of IBI.

### 4.3 Exploration of Potential Mechanisms

Neutrophils secrete antimicrobial proteins and decondensed chromatin DNA, which collectively create extracellular fibrous networks termed Neutrophil Extracellular Traps (NETs)^38^. Hyperglycemia can induce the release of circulating NET biomarkers, and serum NET levels in patients correlate strongly with their concomitant renal and cardiovascular diseases^39^. Endothelial dysfunction can be directly caused by NETs, which can then promote the development of atherosclerosis^40^. Furthermore, neutrophils participate in the inflammatory activation process in kidney diseases, promoting the production of pro-fibrotic cytokines and growth factors, leading to renal fibrosis^41^. T lymphocytes are key regulators in atherosclerosis formation. Regulatory T cells (Tregs) suppress atherosclerosis by enhancing efferocytosis and stimulating the production of pro-resolving mediators that promote atherosclerosis resolution^42,43^. Using mouse and porcine myocardial infarction models, researchers discovered that following an acute myocardial infarction, CD8⁺ T lymphocytes migrate to ischemic cardiac tissue, become activated, and release granzyme B, which leads to apoptosis of cardiomyocyte, adverse ventricular remodeling, and a decline in myocardial function. Additionally, depleting CD8⁺ T lymphocytes decreased apoptosis in the ischemic myocardium, reduced inflammation, minimized myocardial damage, and enhanced cardiac function^44^. A Mendelian randomization study suggested that increased absolute counts of CD8⁺ T cells and CD4⁺CD8dim T cells may elevate the risk of type 2 diabetes^45^. The advancement of chronic kidney disease is also closely associated with lymphocytes. Jiang et al., utilizing single-cell RNA sequencing and other methods, discovered that T cells play a crucial role in the transition from acute kidney injury (AKI) to CKD. CD8⁺ T cells induce endothelial cell apoptosis via the Fas ligand (FasL) - Fas signaling pathway, leading to peritubular capillary rarefaction and renal fibrosis^46^.

The systemic inflammatory state represented by IBI can directly promote atherosclerosis. Neutrophils impair endothelial function by releasing neutrophil extracellular traps (NETs), while the imbalance of lymphocyte subsets (e.g., CD8⁺ T cells) contributes to plaque instability and myocardial injury. The “threshold effect” observed in our study may correspond to the critical point at which inflammatory activation triggers plaque rupture or acute coronary events. Inflammation is a core link in the development of insulin resistance and diabetes mellitus^45^. Meanwhile, as a target organ of inflammation, an elevated IBI may reflect the progressive renal fibrosis process driven by inflammatory mediators (e.g., transforming growth factor-β1, TGF-β1)^36,41,46^. This explains why the risk effect of IBI is more prominent in the subgroup of patients with comorbid metabolic syndrome and CKD.

The robust predictive value of IBI demonstrated in this study stems from the core role of its components in the pathophysiology of CKM syndrome. IBI is not an abstract index, but rather a quantifier of “metabolic inflammation”—a key driver of the syndrome. As an acute-phase protein produced by the liver, C-reactive protein (CRP) levels directly reflect the systemic activity of core inflammatory cytokines such as interleukin-6 (IL-6); the IL-6 signaling pathway has been confirmed to be directly involved in the progression of insulin resistance, atherosclerosis, and heart failure^47–49^. Neutrophils, as the rapid-response force of innate immunity, are not only indicative of vascular endothelial injury caused by NET formation^38,40^, but also closely associated with adipose tissue inflammation and systemic insulin resistance. The balance between lymphocytes, particularly regulatory T cells (Treg) and effector T cells (e.g., CD8⁺ T cells), is critical for maintaining immune homeostasis; in CKM syndrome, this balance is disrupted and shifts toward a pro-inflammatory state, accelerating atherosclerosis and end-organ fibrosis^42,46^. Therefore, by integrating these three dimensions, IBI may better capture the complex and persistent inflammatory state in CKM syndrome than any single marker—this explains the robustness of IBI as an independent predictor.

Both all-cause mortality and cardiovascular mortality exhibited significant non-linear relationships with the Inflammatory Burden Index (IBI) in the CKM syndrome population. All-cause mortality: The restricted cubic spline (RCS) curve for IBI and all-cause mortality revealed a nadir (point of lowest risk) at IBI = 3.357 (HR = 0.479, 95% CI: 0.251–0.914). The curve intersected the HR=1 reference line at two points (IBI = 0.0864 and IBI = 52.49), defining an “optimal inflammatory range” (0.0864–52.49). This suggests that for CKM syndrome patients, maintaining the inflammatory burden within this range, particularly near the nadir, may confer a survival advantage. Deviating from this range—whether too low or too high—increases all-cause mortality risk. All-cause mortality encompasses deaths from various causes (cardiovascular disease, respiratory diseases, infections, renal failure, etc.). An extremely low inflammatory state may reflect immunosuppression, malnutrition, or severe chronic wasting diseases, increasing susceptibility to infections, poor wound healing, and consequently, mortality risk. Conversely, chronic long-term inflammation contributes to the development of numerous diseases (e.g., neurodegenerative disorders, cancer, cardiovascular diseases) and also elevates mortality risk^50^. Although chronic inflammation is mainly investigated in the context of disease progression, it often starts as an adaptive response to stress, much like acute inflammation. In the context of acute infection, it is thought that insulin resistance facilitates the mobilization of nutrients to bolster immune system activation, thereby meeting the heightened energy demands^51,52^. Inflammatory mechanisms may also have developed to react to non-infectious or non-injury stressors, such as metabolic and physical stress. For instance, obesity leads to inflammation in adipose tissue, which consequently causes insulin resistance; this development of insulin resistance may initially serve a beneficial purpose by promoting the mobilization of nutrients from adipose tissue to help mitigate obesity^53^. Therefore, maintaining the inflammatory burden within a specific range may minimize all-cause mortality risk. Cardiovascular mortality: The RCS curve for IBI and cardiovascular mortality displayed an inflection point at IBI = 1.684 (HR = 0.764, 95% CI: 0.26–2.24). Left of the inflection point (Low-risk region): Risk changes were relatively modest. This may correspond to lower levels of metabolic inflammation, minimal or compensated cardiovascular damage, resulting in a relatively stable cardiovascular mortality risk close to baseline. To the right of the inflection point, the slope of the curve increased significantly, suggesting a steep rise in the risk of cardiovascular mortality. This steep increase likely reflects acute events triggered by inflammation, such as plaque rupture, severe endothelial dysfunction, or malignant arrhythmias. For example, during myocardial infarction, necrotic cardiomyocytes release damage-associated molecular patterns (DAMPs) that interact with pattern recognition receptors (e.g., Toll-like receptors). Together with complement activation and reactive oxygen species (ROS), these induce the upregulation of cytokines and chemokines, promoting both coronary and systemic inflammatory responses^54,55^. This inflammatory process can further impair the myocardium, triggering heart failure (HF) events and propagating extensive inflammation across the entire coronary tree^56^. Furthermore, cholesterol crystals activate the pyrin domain of NOD-like receptor family containing 3 (NLRP3) inflammasome, which is followed by the activation of the interleukin-1β (IL-1β) – C-reactive protein (CRP) and the inflammatory cascade. Both local coronary and systemic inflammation can promote atherothrombosis and respond to cardiomyocyte death within the necrotic myocardium^57^.

### 4.4 Clinical and Public Health Implications

Based on routine and low-cost hematological test parameters, IBI is promising to serve as a practical risk stratification tool in primary care settings and resource-limited environments. It can help identify CKM patients who, despite adequate control of traditional risk factors, remain at high risk due to elevated inflammatory burden. The identified “U-shaped” relationship suggests that a lower inflammatory burden is not necessarily better. Future studies need to explore whether there exists an individualized “inflammatory target value”. Additionally, IBI may be used to identify specific patient subgroups that are likely to benefit from emerging anti-inflammatory therapies (e.g., interleukin-1 β (IL-1 β) inhibitors). Our findings, particularly the “U-shaped” relationship between IBI and mortality, offer profound and novel implications for clinical practice. First, they suggest that the “lower is better” paradigm in inflammation management may not be fully applicable to CKM syndrome. There exists a potential “optimal inflammation window,” where both excessively high and excessively low inflammatory burden may be harmful. This calls for future studies to explore individualized inflammatory target values, rather than blindly pursuing the minimization of inflammatory markers. Second, the simplicity of IBI makes it highly suitable for dynamic risk monitoring. Future research could investigate whether changes in IBI during follow-up have greater prognostic value than a single measurement, thereby identifying patients at high risk of “inflammatory progression.” Most importantly, our study provides epidemiological evidence for targeted anti-inflammatory therapy. For instance, could patients with high IBI and in the advanced stages of CKM syndrome be prioritized as beneficiaries of drugs such as interleukin-6 (IL-6) inhibitors, sodium-glucose cotransporter 2 (SGLT2) inhibitors (known to have anti-inflammatory effects), or colchicine? IBI may be used for patient selection in future clinical trials to verify whether “inflammation-targeted” treatment strategies can improve hard endpoints in CKM syndrome.

Study Strengths: (1)Extensive Cohort & Innovative Focus: This research involves a large-scale cohort analysis that methodically examines the relationship between the Inflammatory Burden Index (IBI) and long-term negative outcomes in patients with CKM syndrome, offering an in-depth exploration of the nature of this association. This innovative approach not only addresses a significant research gap in this field but also offers crucial reference data for future studies. (2) Methodological Advancements: Substantial improvements have been achieved compared to earlier studies by employing the Predicting Risk of CVD Events (PREVENT) equations, which have recently been endorsed by the American Heart Association (AHA) for the prediction of subclinical cardiovascular disease (CVD). The novel race-free

Chronic Kidney Disease Epidemiology Collaboration (CKD-EPI) 2021 equation was applied to estimate the glomerular filtration rate (eGFR). These methodological improvements significantly enhance the reliability of our results. (3) Comprehensive Analytical Strategy: IBI was analyzed both as a categorical variable (divided into quartiles) and as a continuous variable to comprehensively assess its association with all-cause and cardiovascular mortality risk. This dual approach facilitates the identification of risk differences across IBI levels and better reflects clinical reality. (4) Clinically Accessible Biomarkers: The components of IBI—C-reactive protein (CRP), neutrophil count, and lymphocyte count—are readily measurable, widely accessible, and relatively low-cost. These characteristics make IBI highly valuable in clinical practice, particularly within primary healthcare settings. (5) Strongity: We used sensitivity analyses, which confirmed our primary findings’validity.

Study Limitations: (1) Potential residual confounding due to unmeasured variables (e.g., respiratory diseases, autoimmune disorders) remains possible. The specific geographic and demographic characteristics of the NHANES sample may restrict the applicability of our findings. Furthermore, focusing solely on IBI without comparing other biomarkers may overlook potentially superior predictors. Even with this drawbacks, this study shows how inflammation affects the condition of CKM syndrome and paves the way for future longitudinal investigations. (2) Although other potential confounders (e.g., respiratory diseases, autoimmune diseases) were not adjusted for in this analysis, a key strength of the NHANES data is its national representativeness, enhancing the credibility of our findings.

## 5 Conclusion

Our study is the first to demonstrate that IBI exhibits significant non-linear associations with both all-cause and cardiovascular mortality in patients with CKM syndrome and serves as a risk factor for these patients. Monitoring the Inflammatory Burden Index holds important clinical significance for assessing mortality risk in individuals with CKM syndrome.

## Abbreviations

IBI: Inflammatory Burden Index
AHA: American Heart Association
BMXBMI: body mass index;
urdratio: urine albumin-to-creatinine ratio;
LBXSUA: serum uric acid;
BPXSY_avg: mean systolic blood pressure;
BPXDI_avg: mean diastolic blood pressure
LBDLDL: low-density lipoprotein;
LBDHDD: high-density lipoprotein;
LBXTC: total cholesterol;
LBXTR: triglyceride;
LBXGH: glycated hemoglobin;
MeTS: metabolic syndrome
CKD: chronic kidney disease;
CKM: cardiorenal metabolic syndrome;
CVD: cardiovascular disease

## Supplementary Information

The supplementary figures and tables in this paper can be found in the supplementary materials.

## Acknowledgements

We extend our sincere appreciation to every participant involved in the NHANES study and the entire project team.

## Author contributions

Qin Yuqi was responsible for study design, conceptualization, data collection, statistical analysis, result interpretation, as well as drafting and revising the manuscript. Gou Luoning was responsible for study design, reviewing and revising the manuscript. All authors read and approved the manuscript.

## Data availability

The datasets generated and analyzed during this research can be retrieved from the NHANES database (https://wwwn.cdc.gov/nchs/nhanes/default.aspx).

## Declarations

Approval from the ethics committee and consent for participation

The research protocol of the NHANES study obtained approval from the Ethics Review Board of the National Center for Health Statistics. Prior to their participation, all subjects signed a written informed consent form.

---

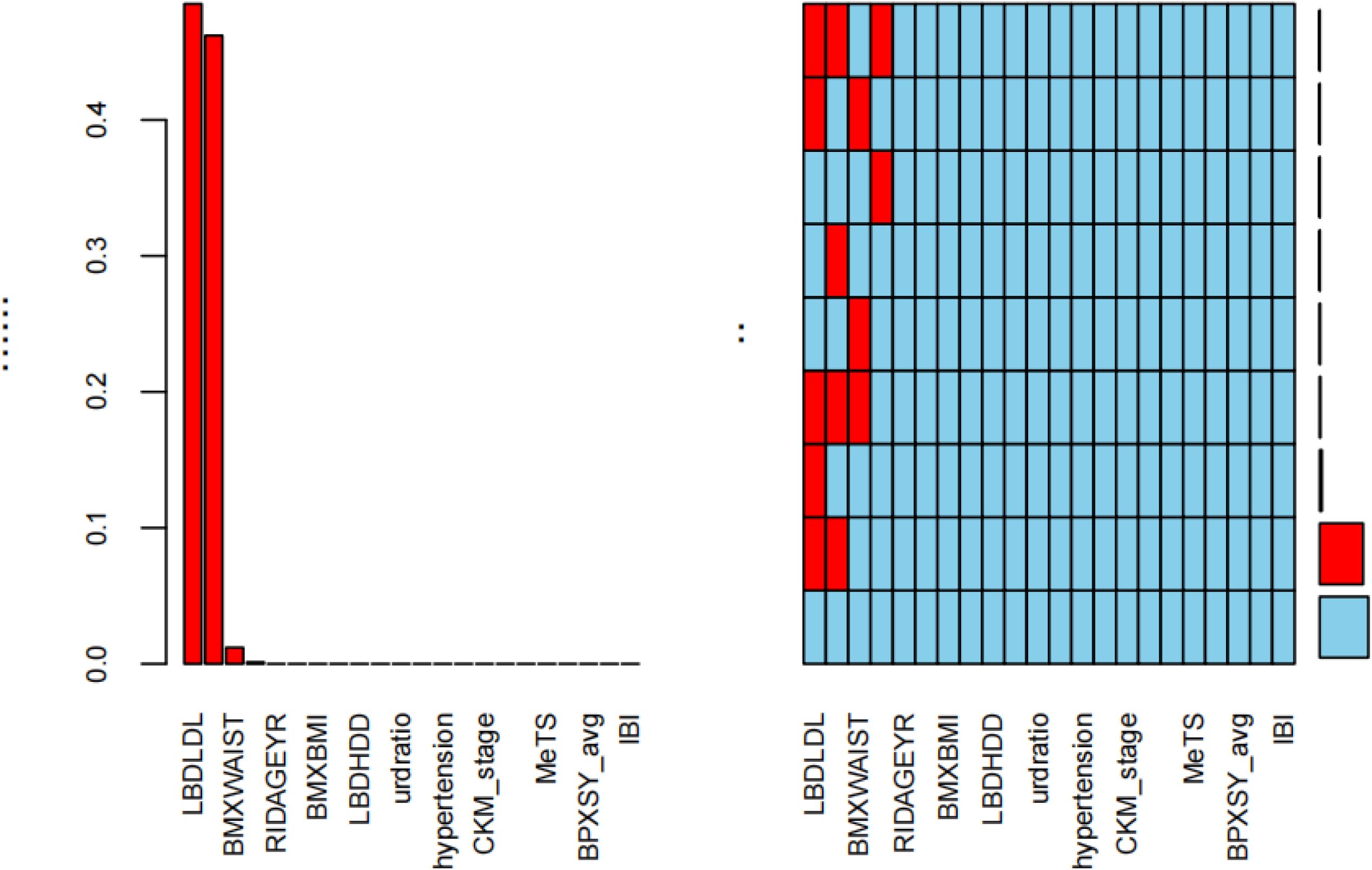

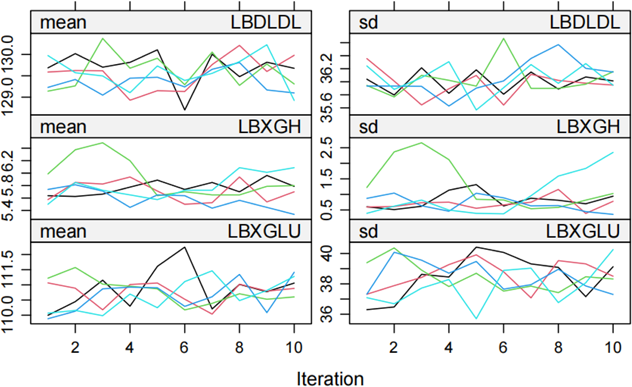

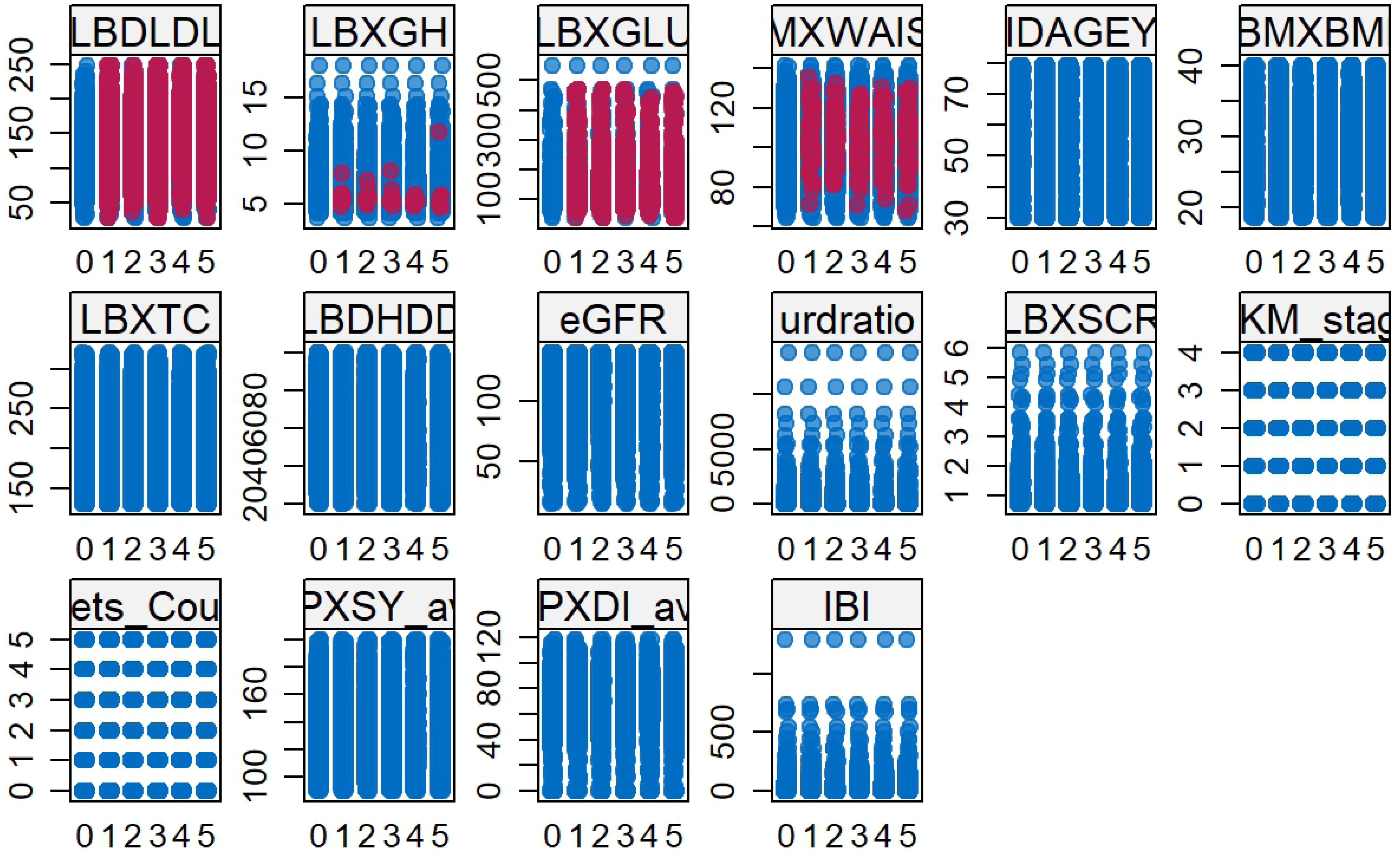

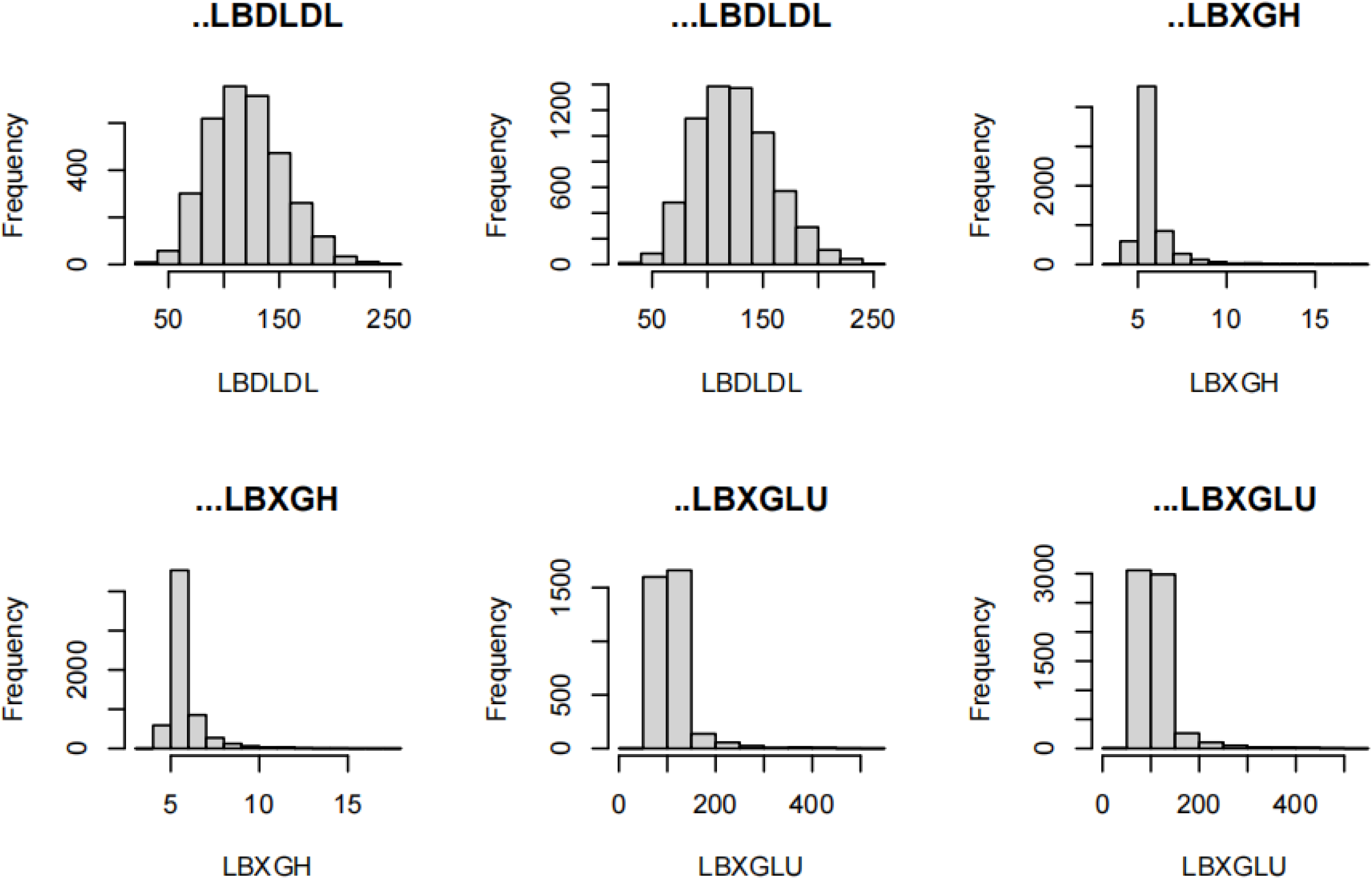

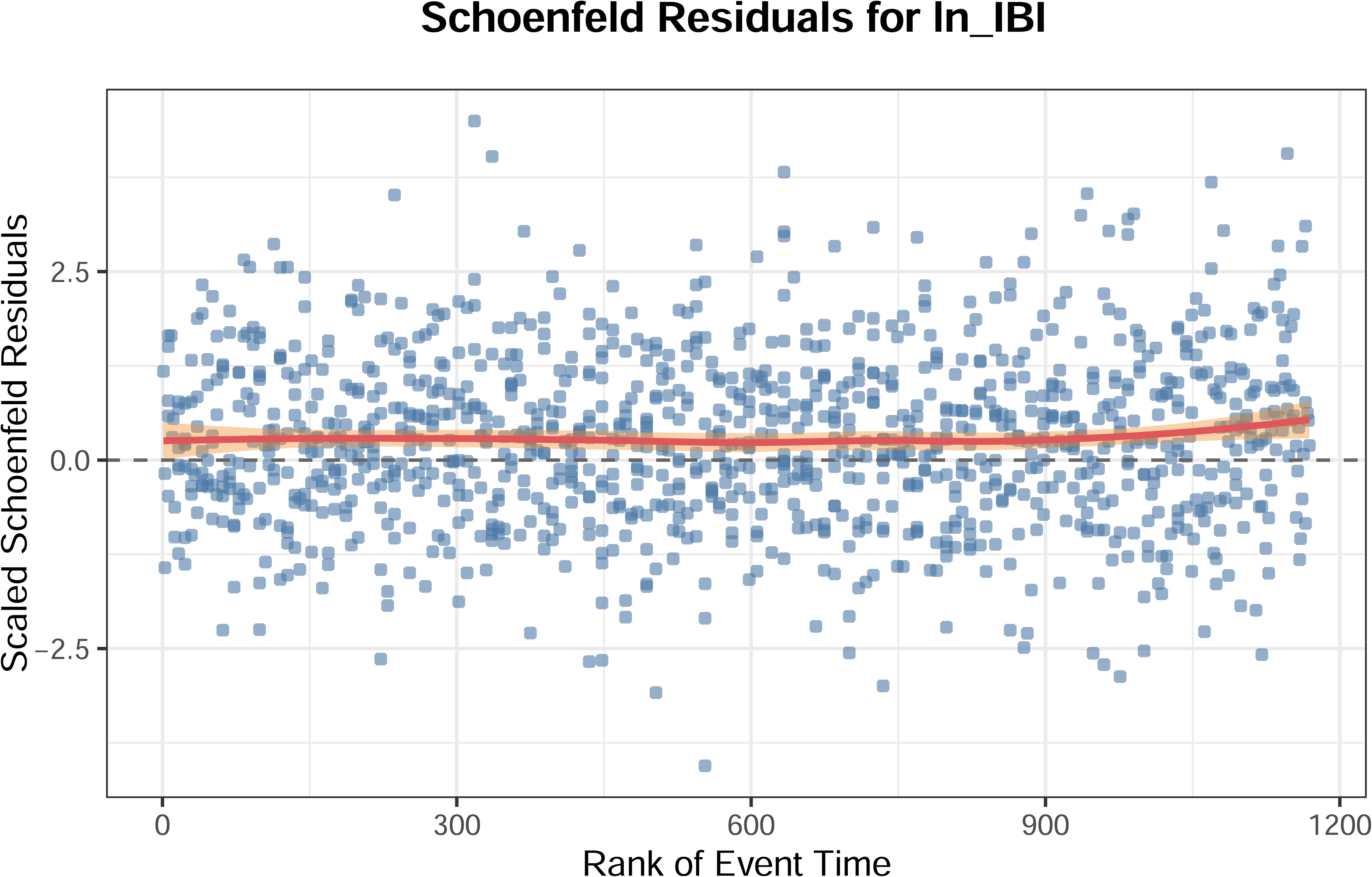

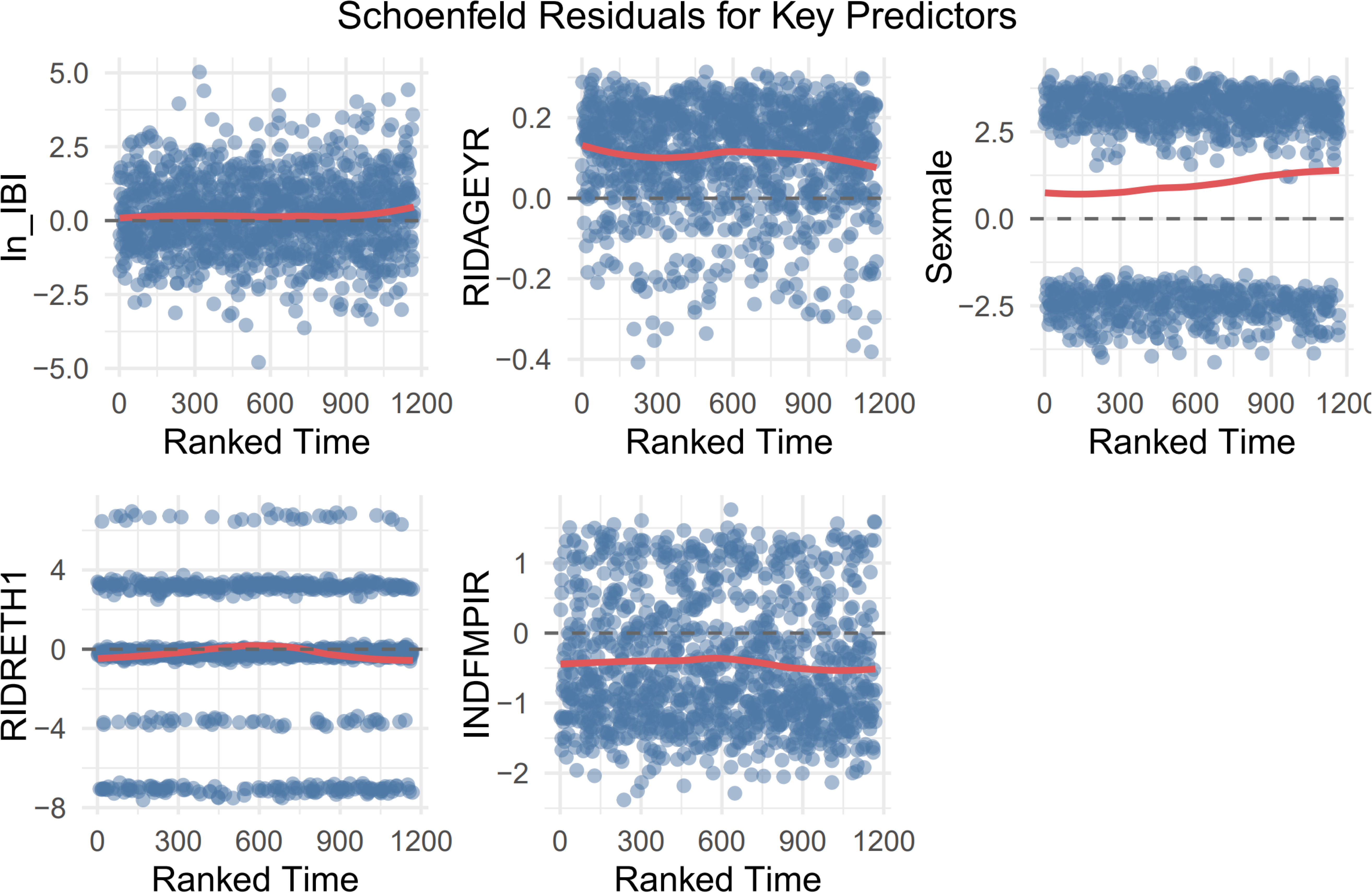

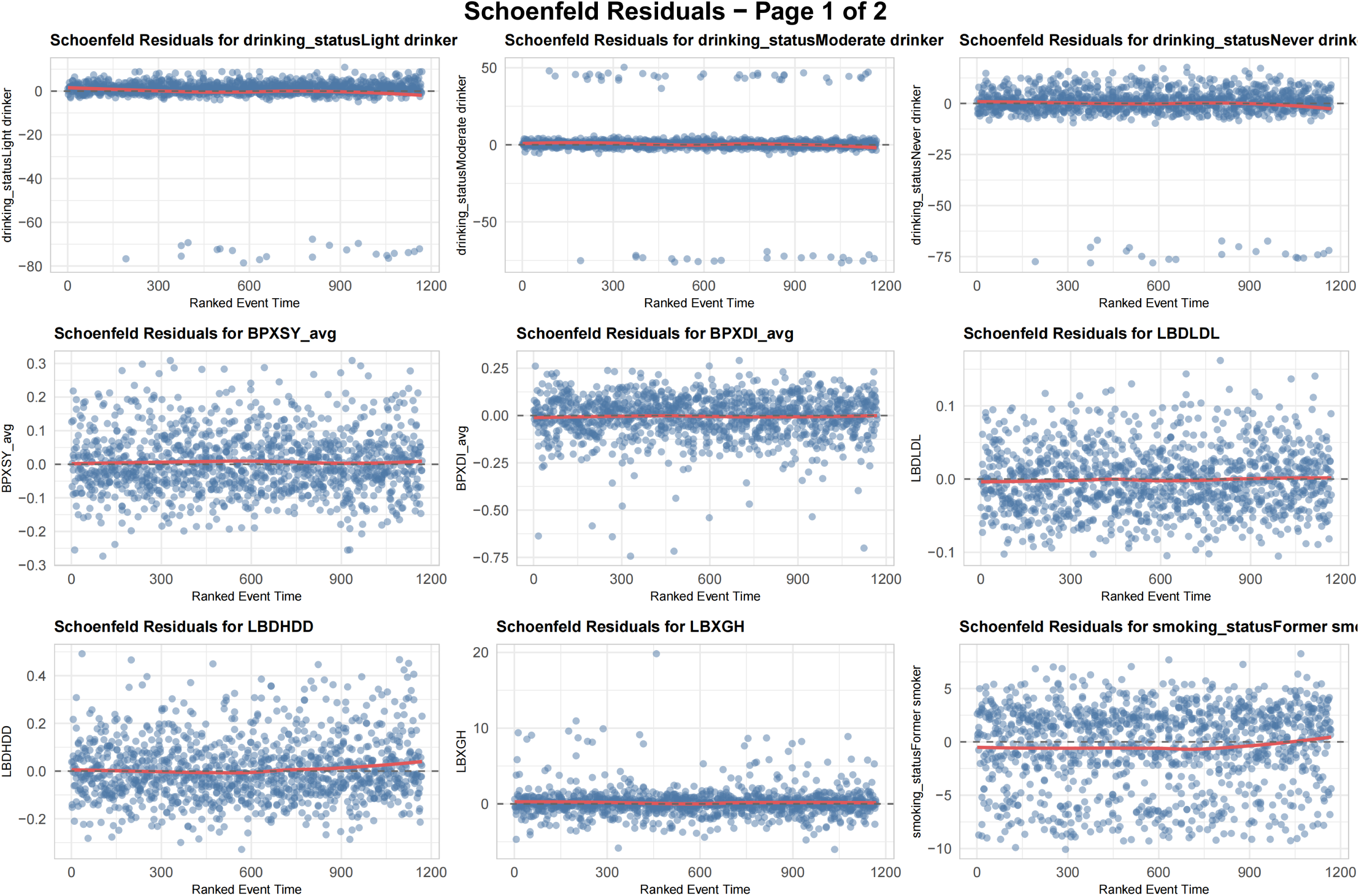

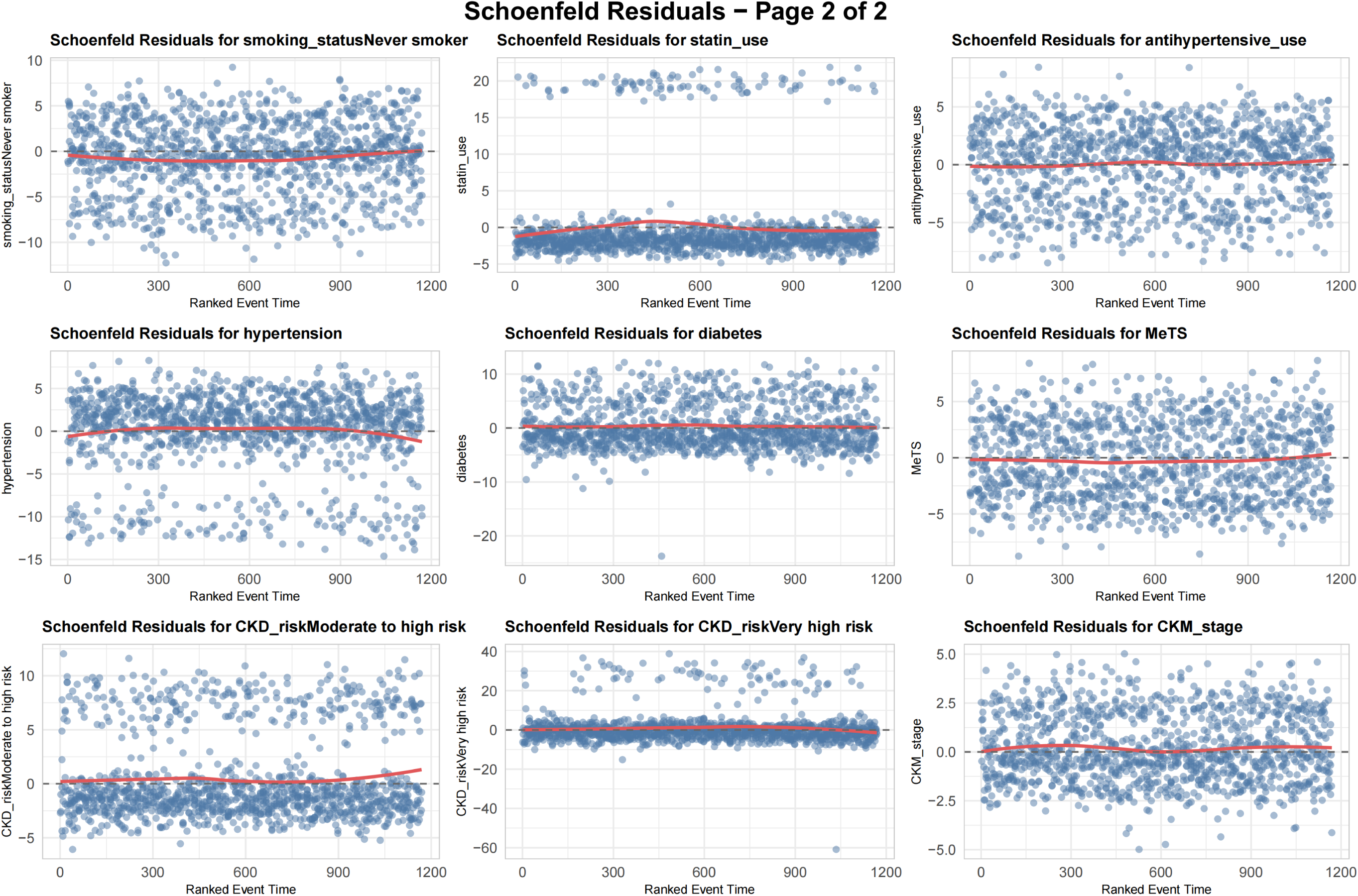

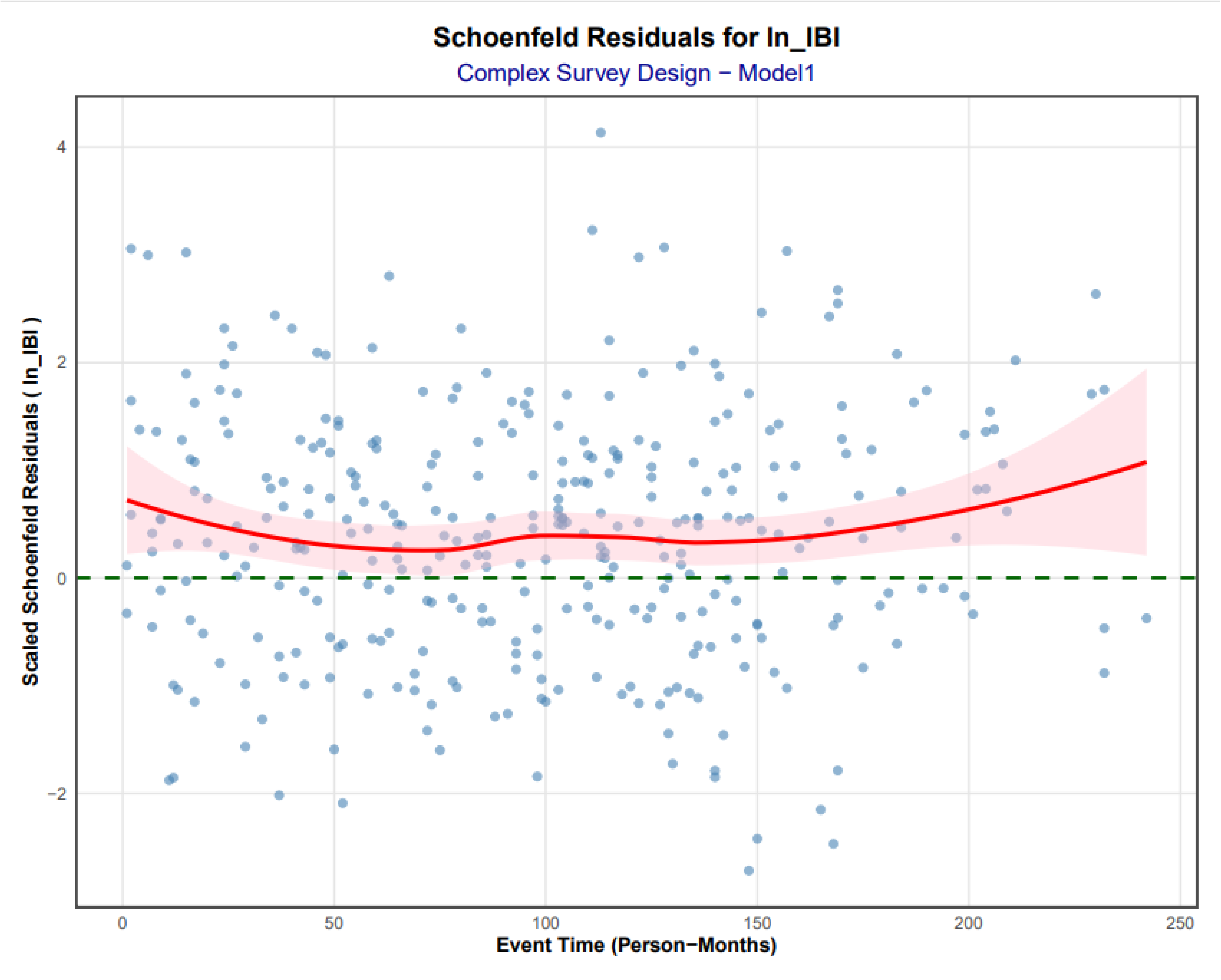

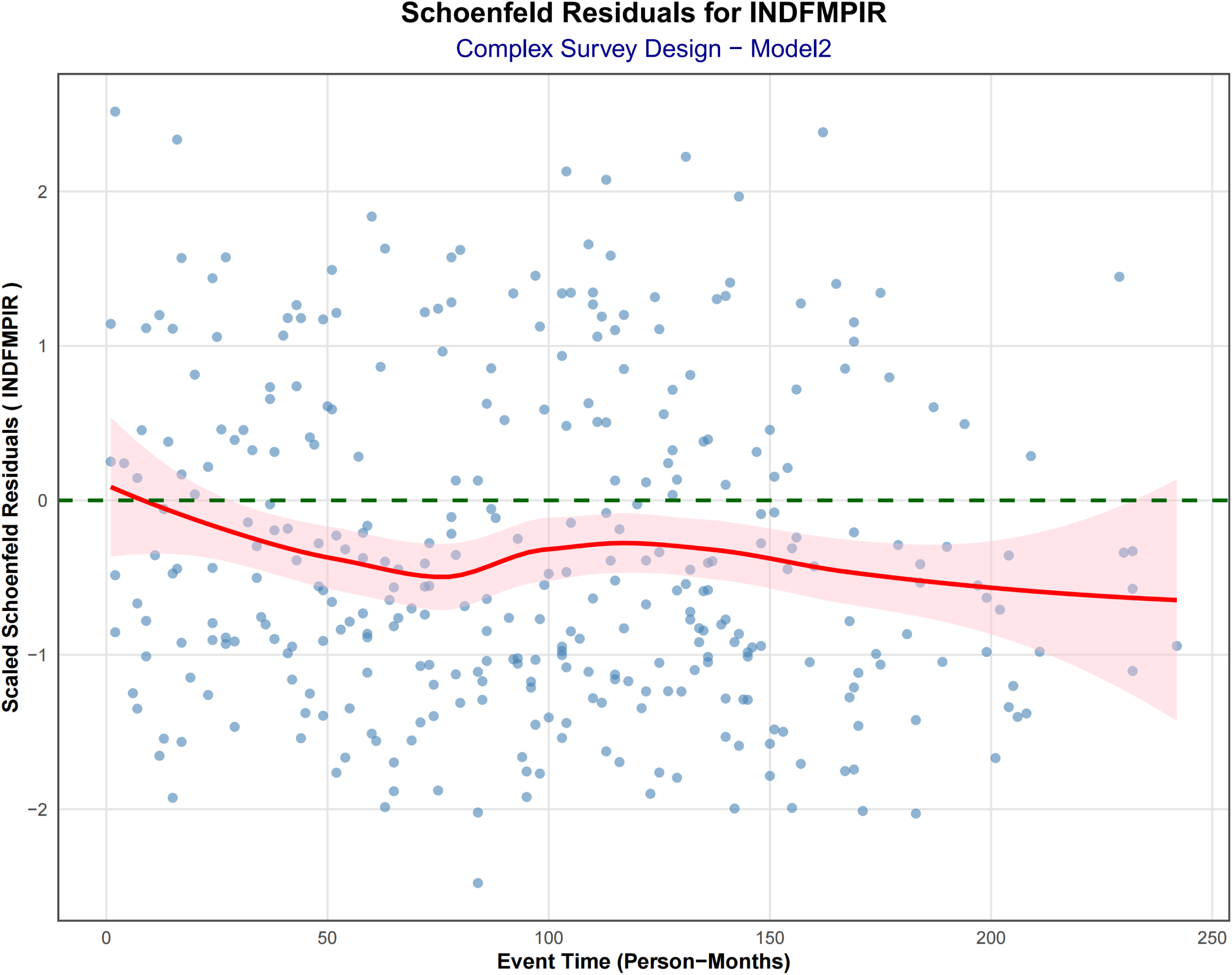

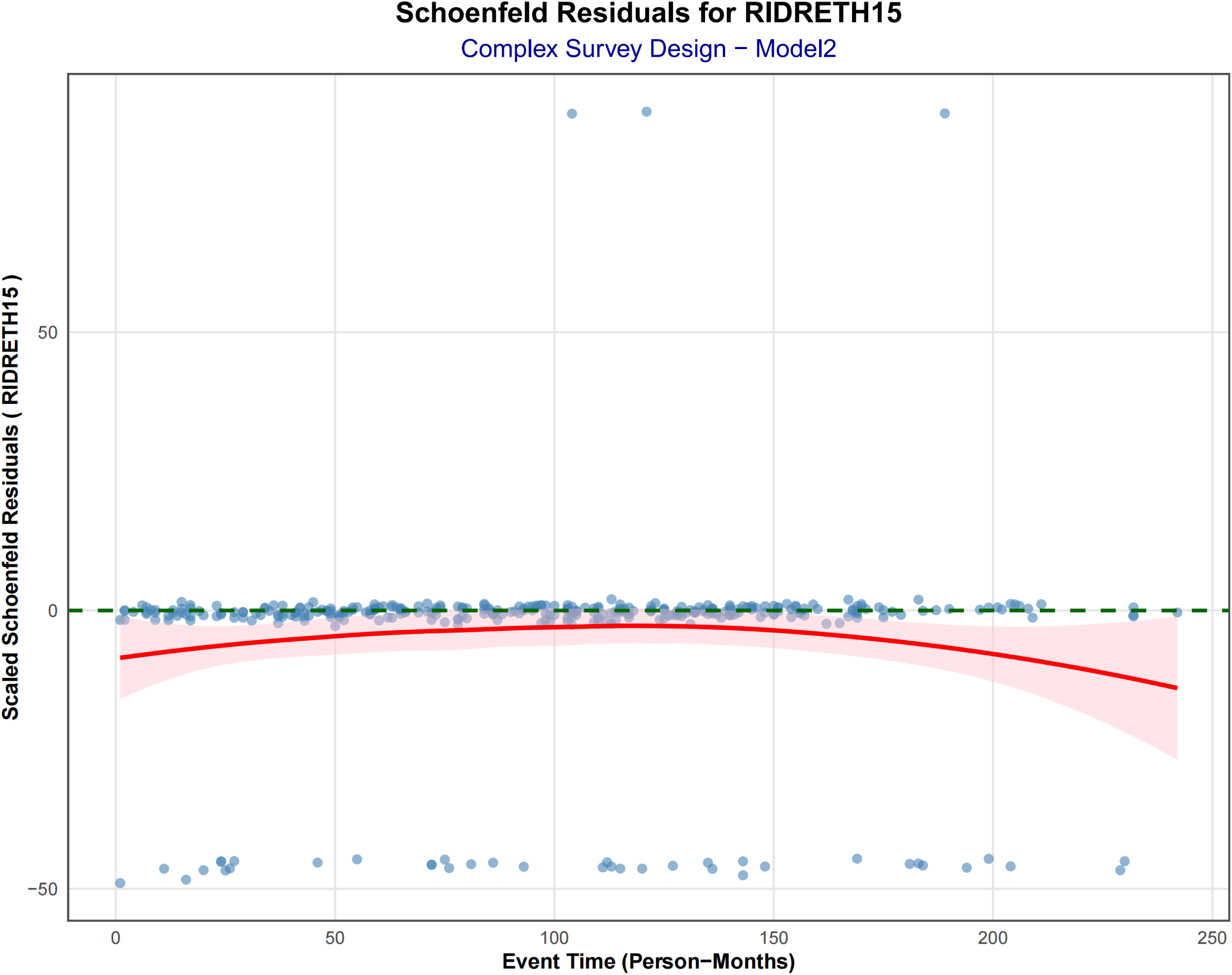

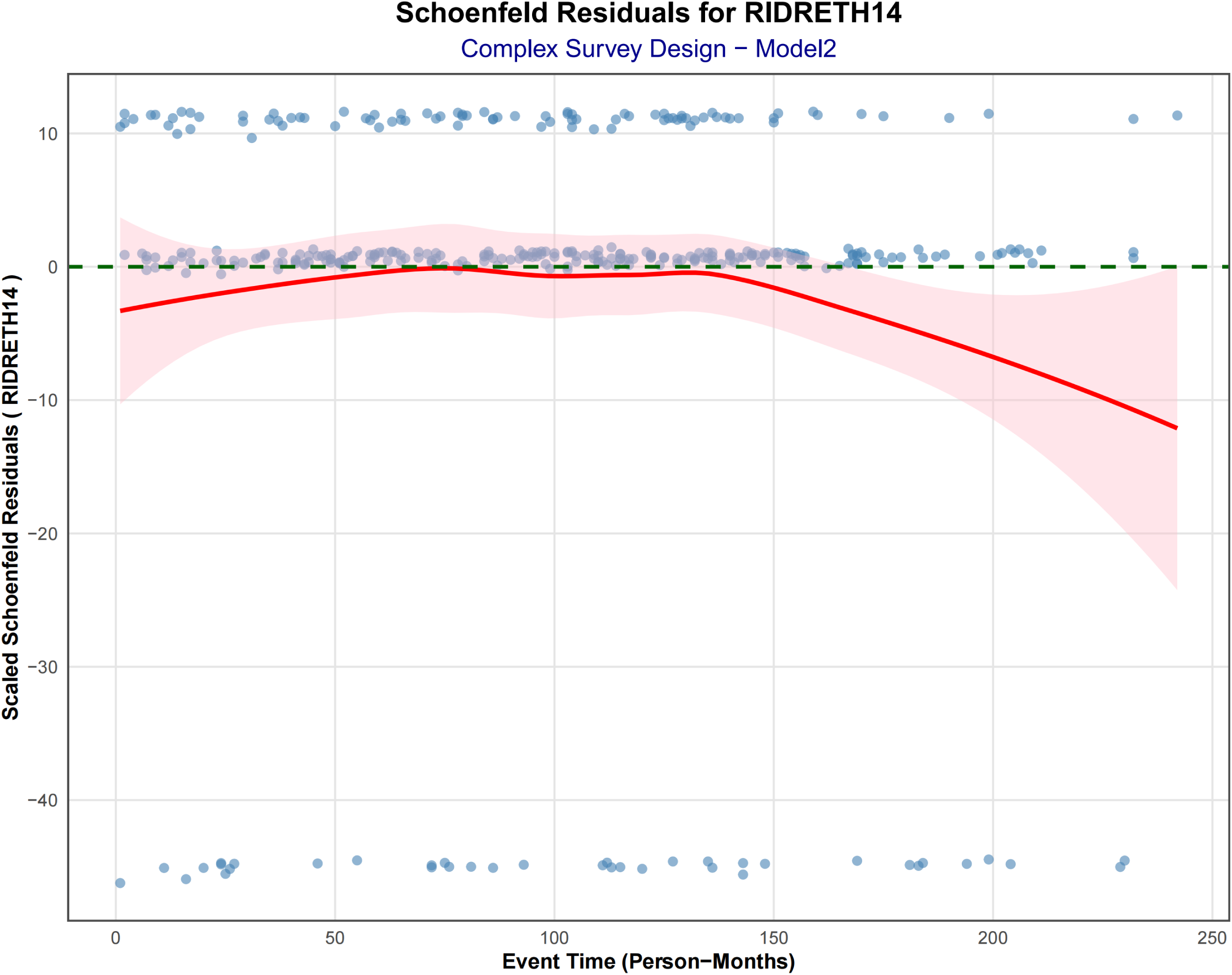

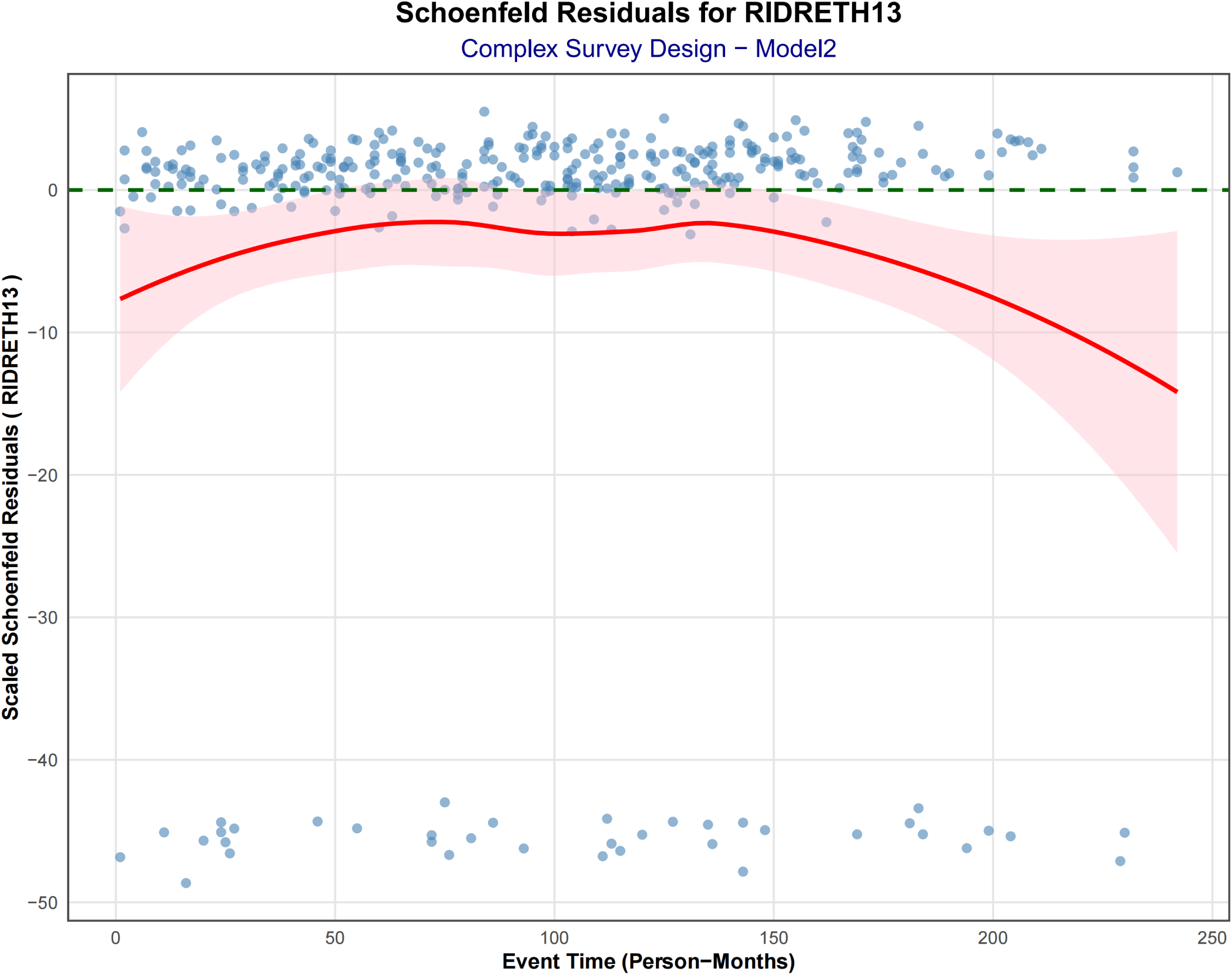

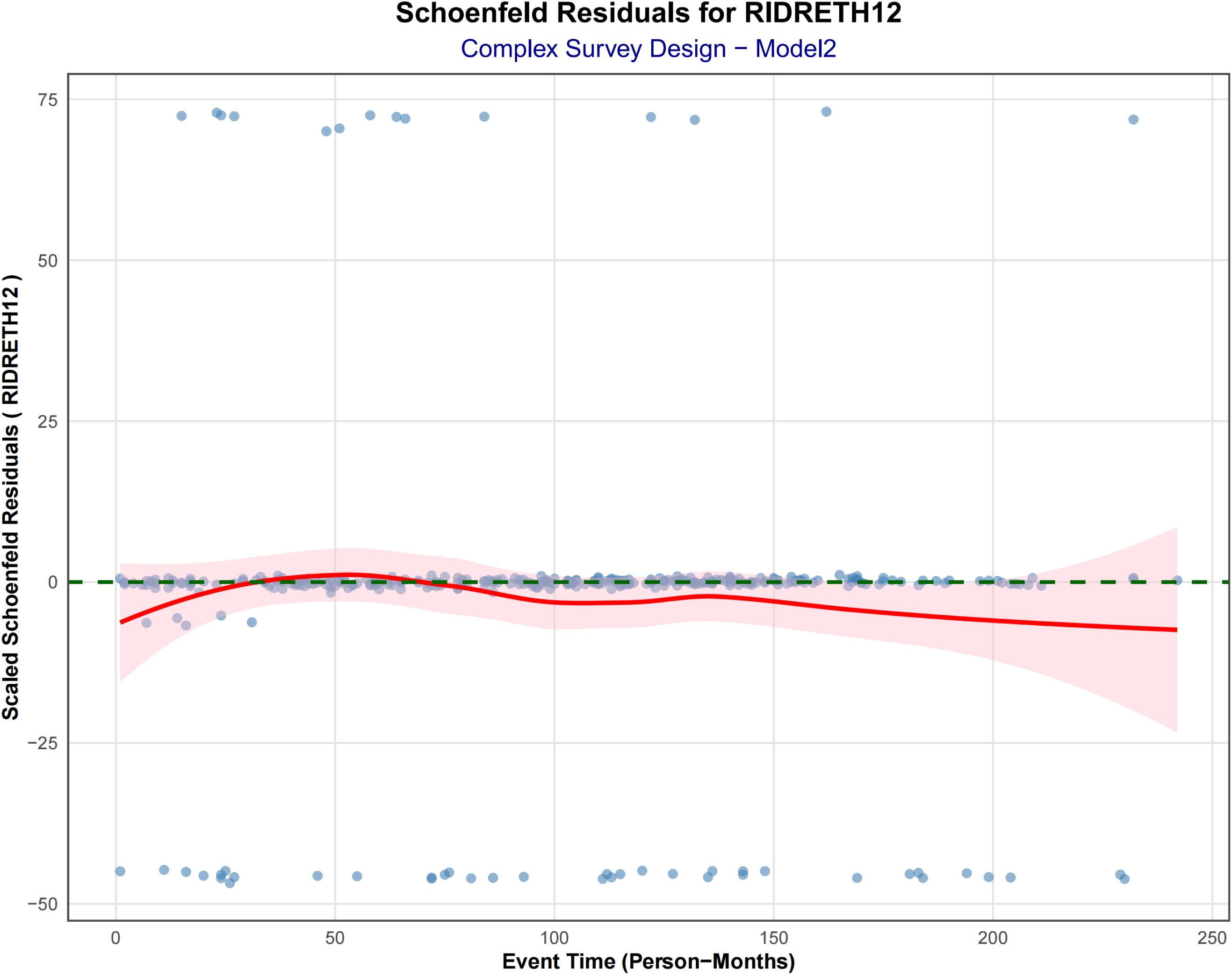

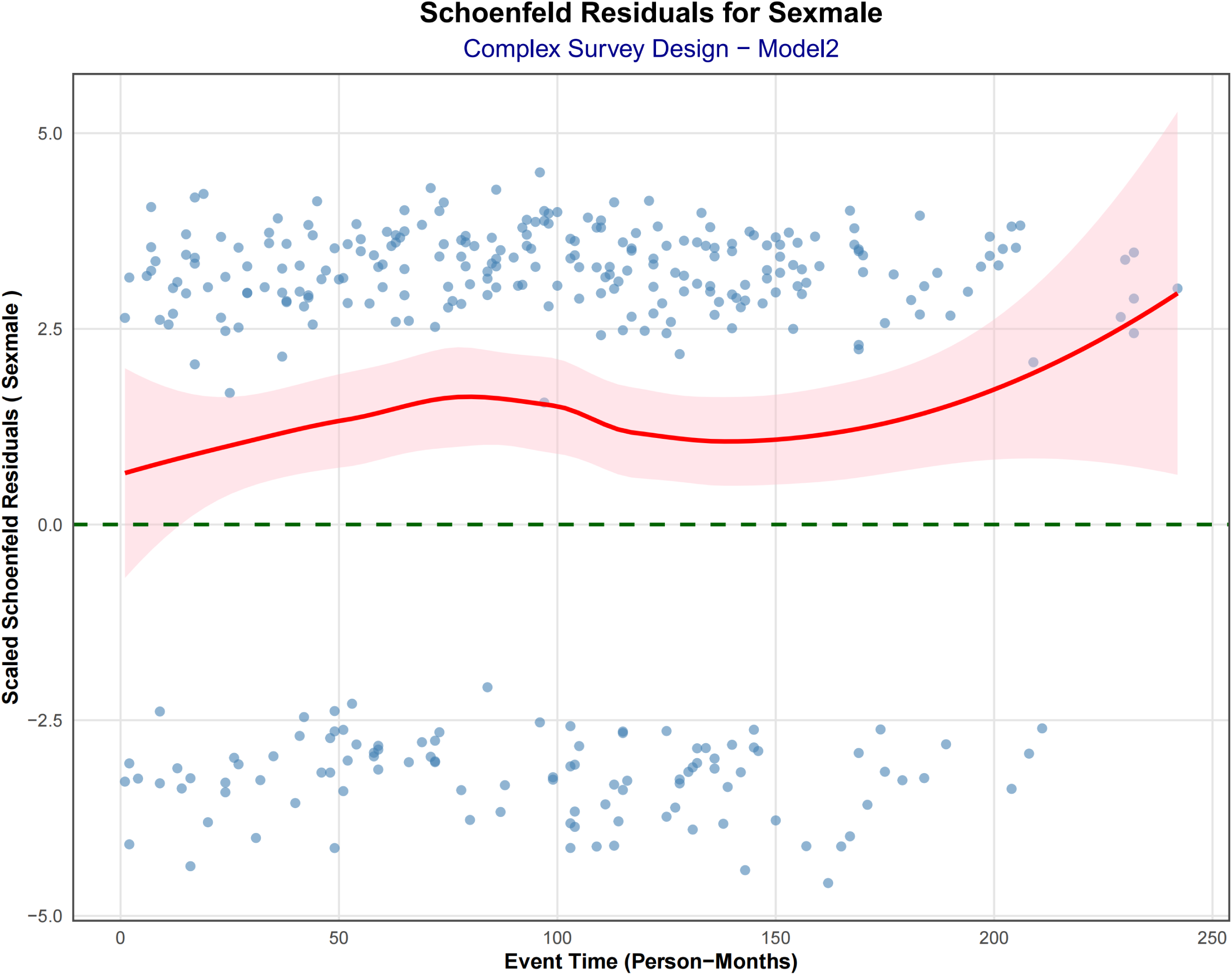

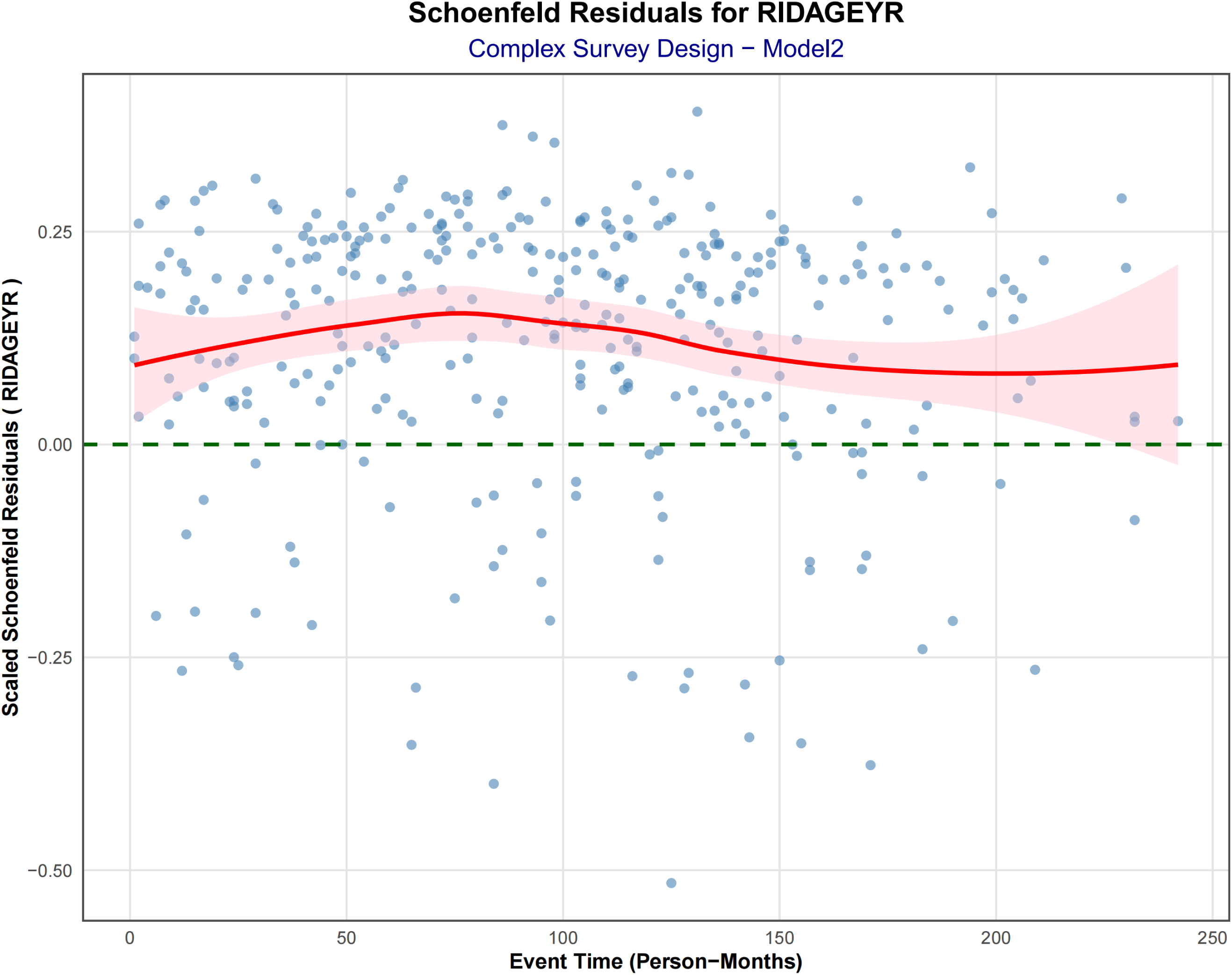

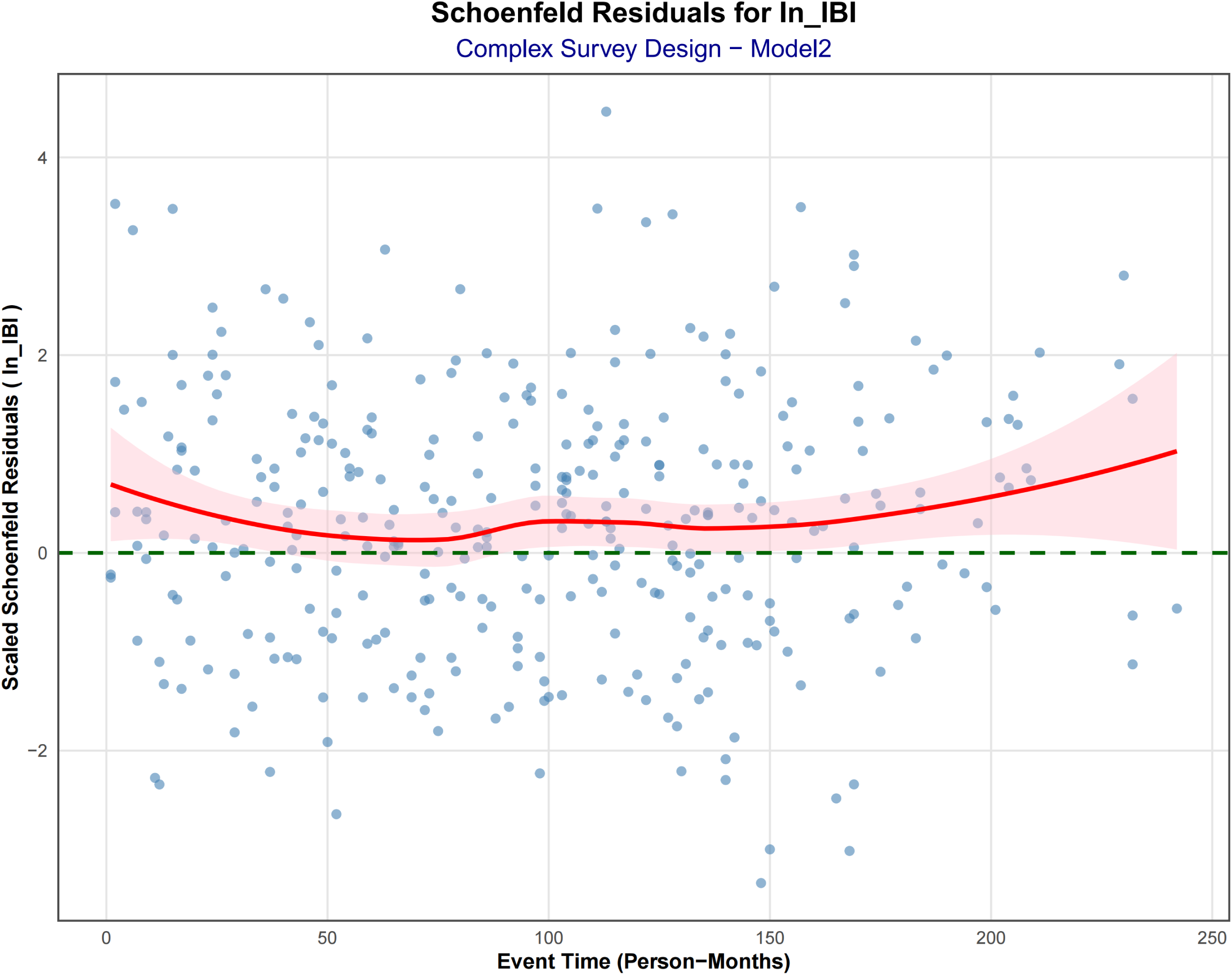

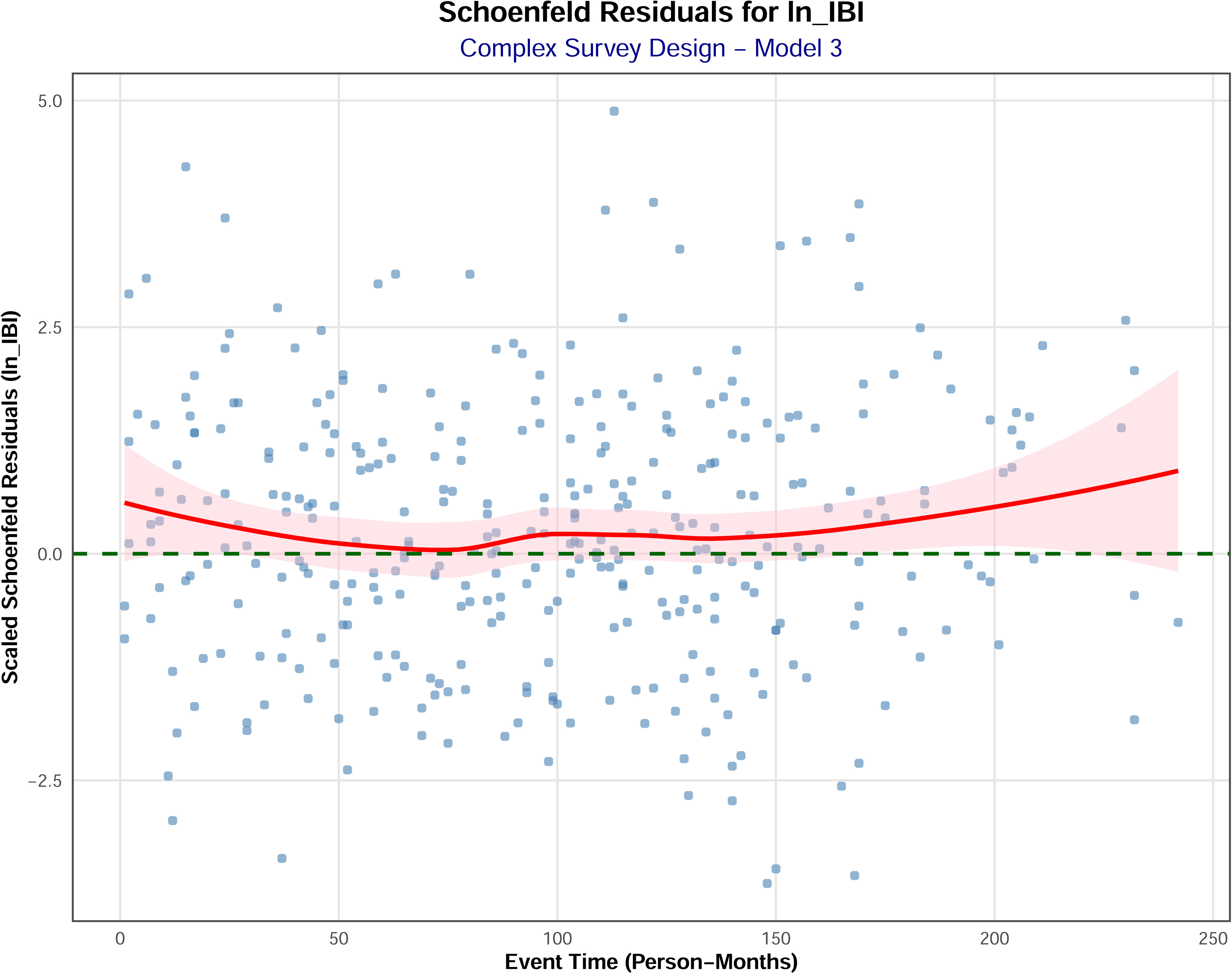

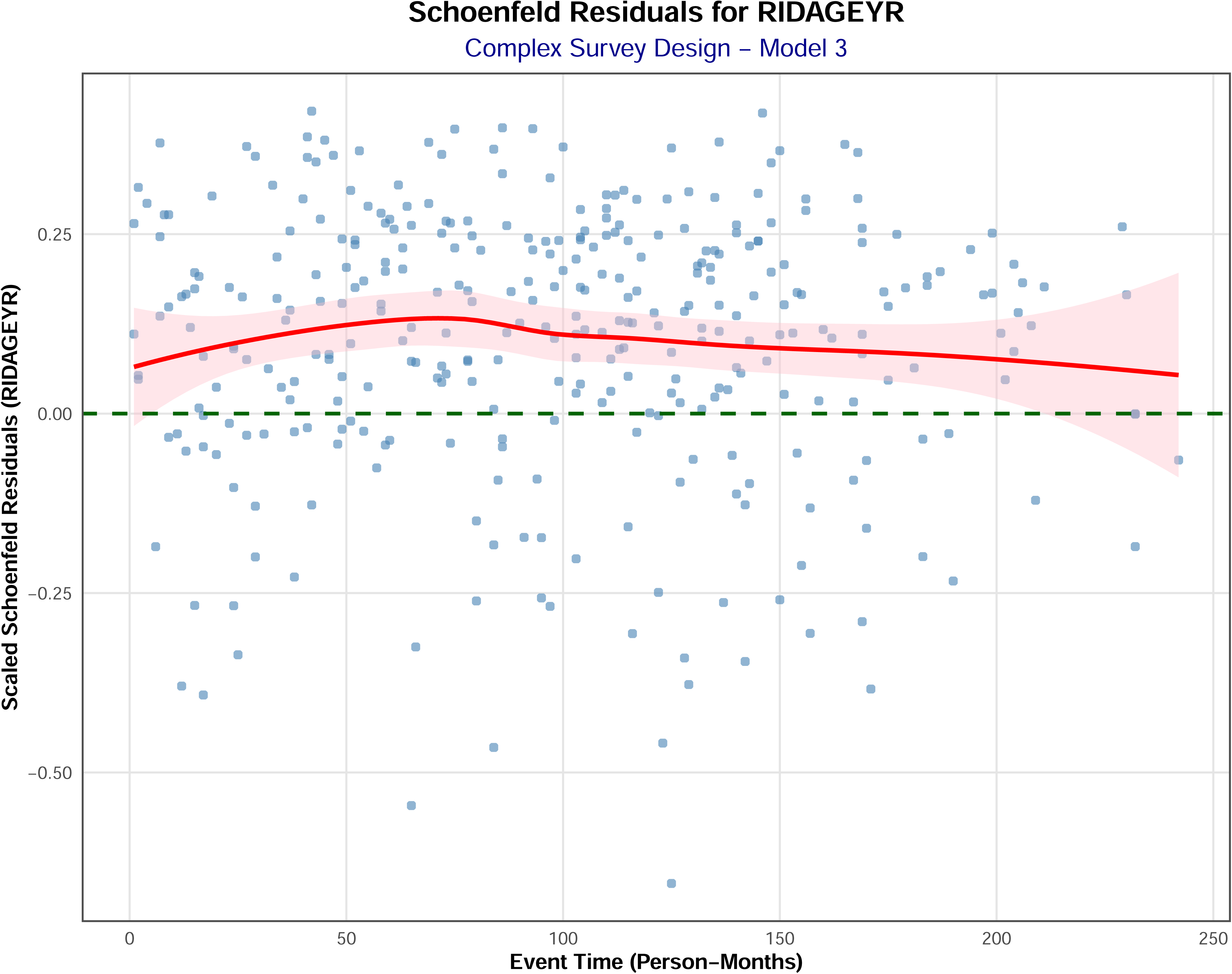

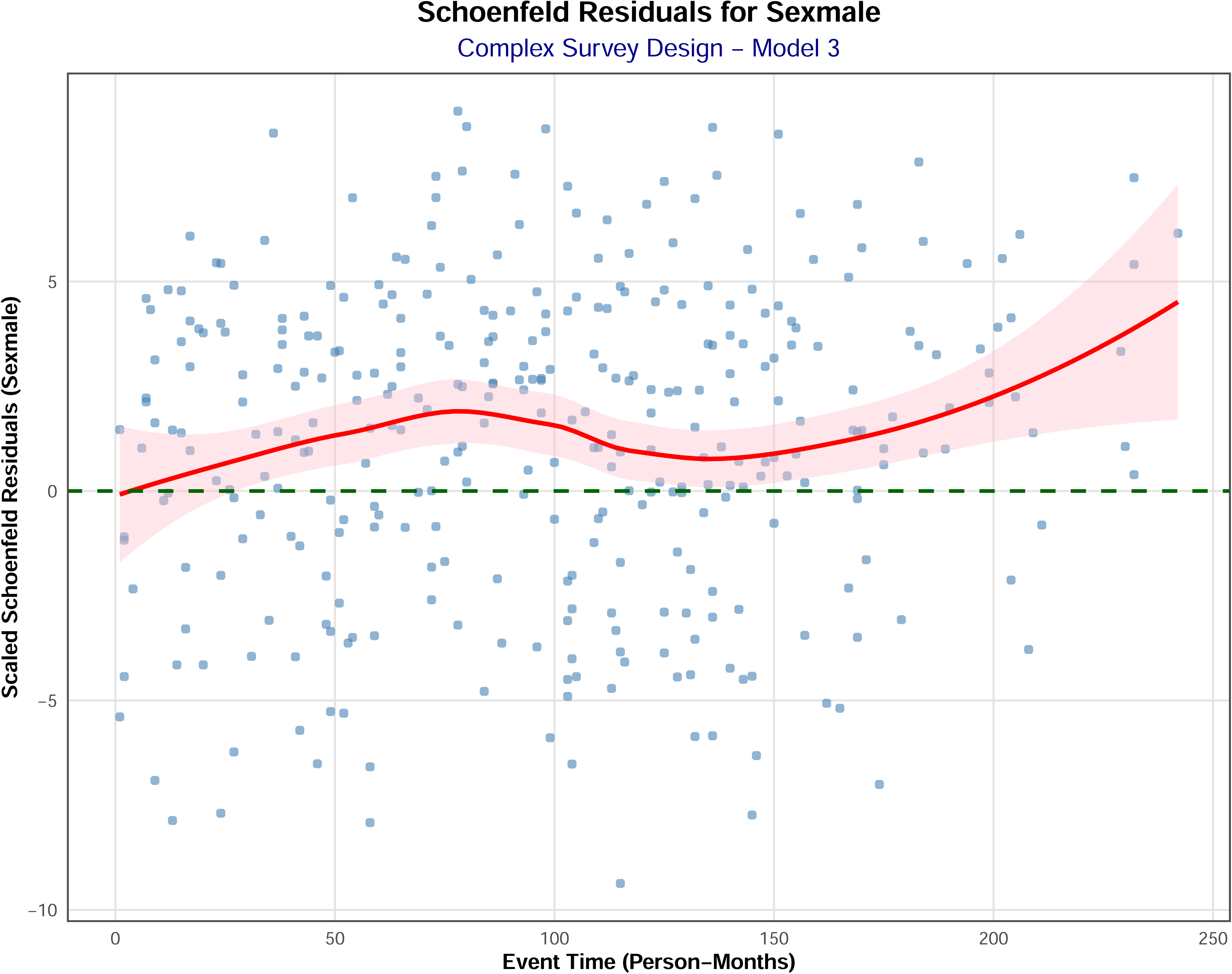

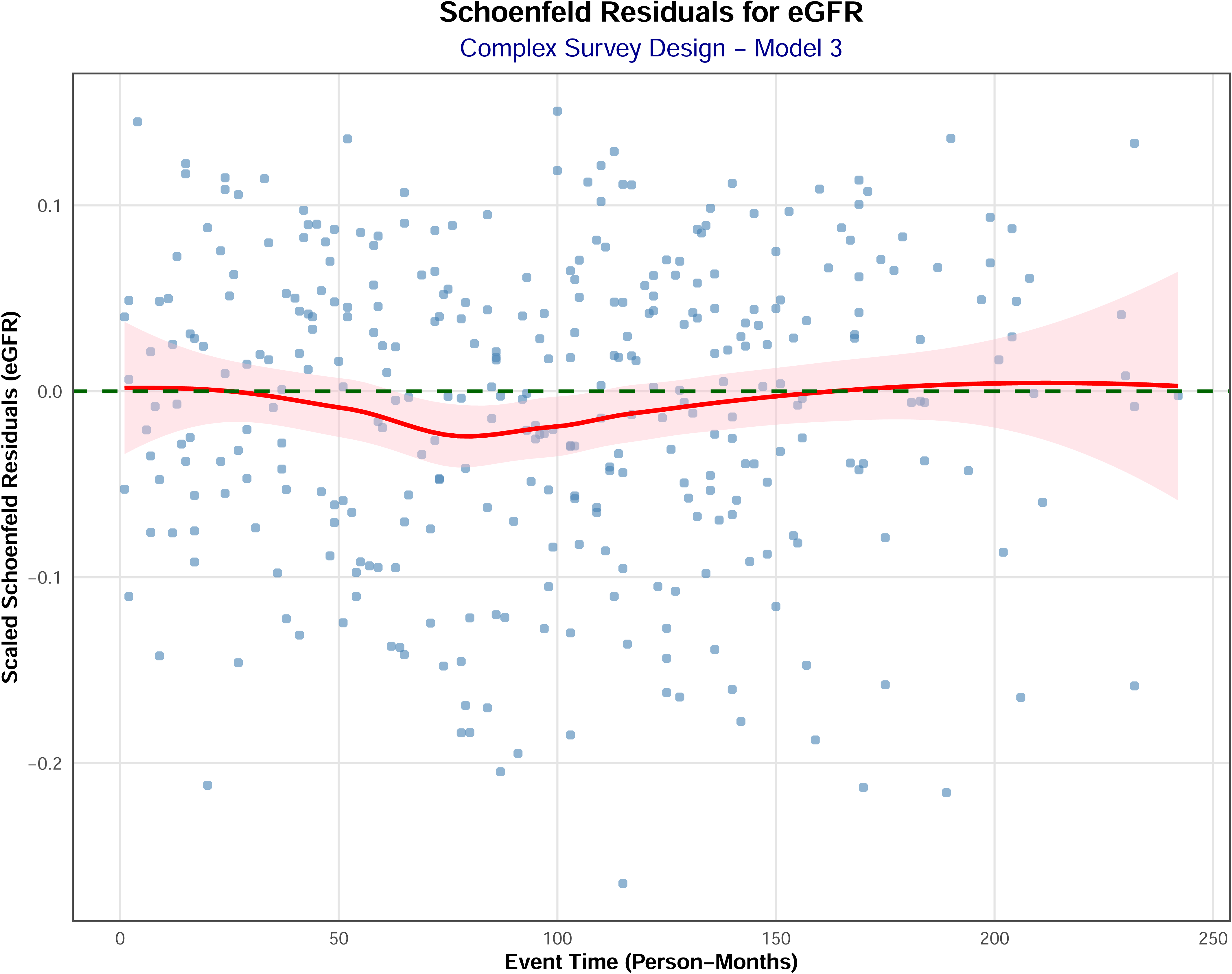

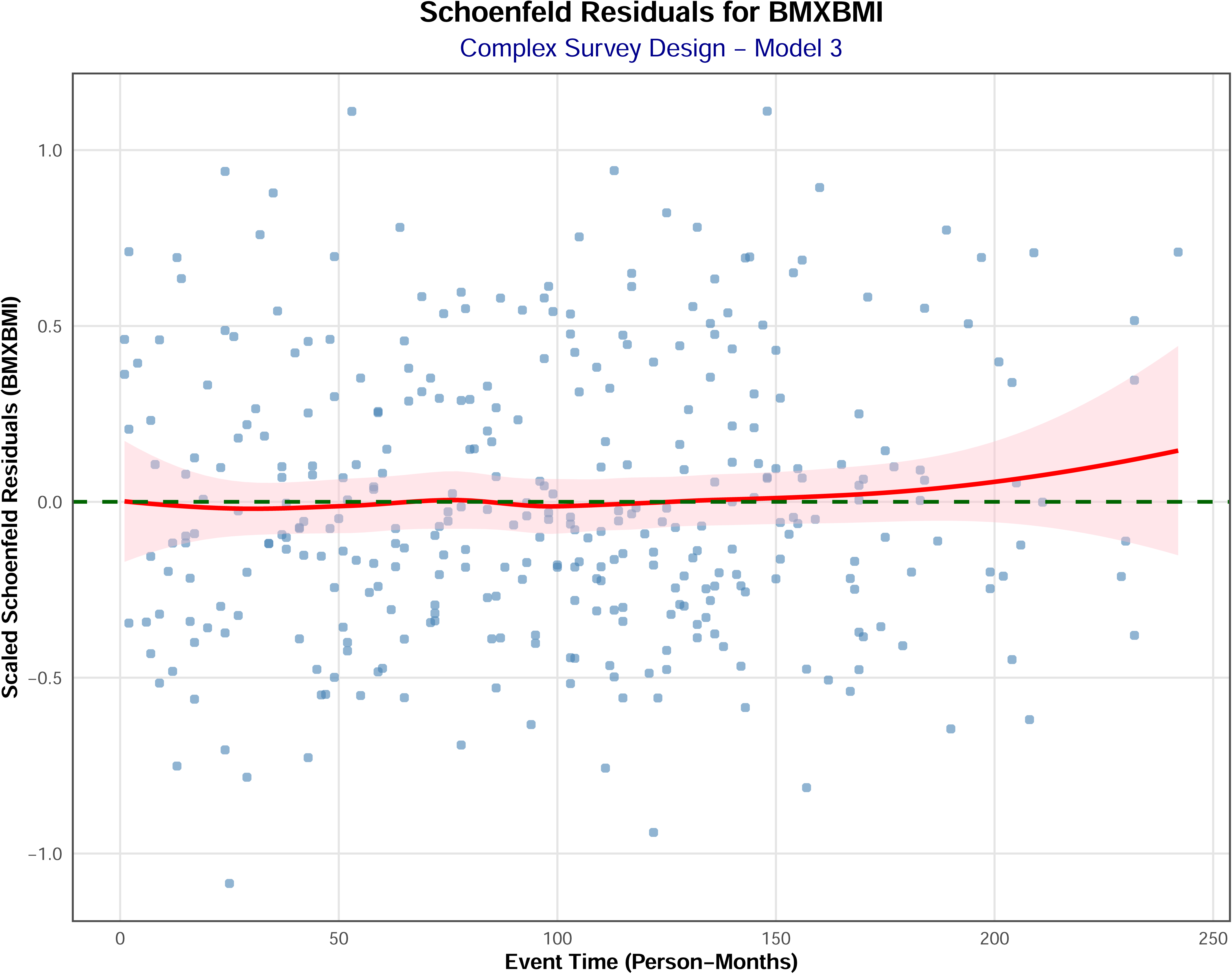

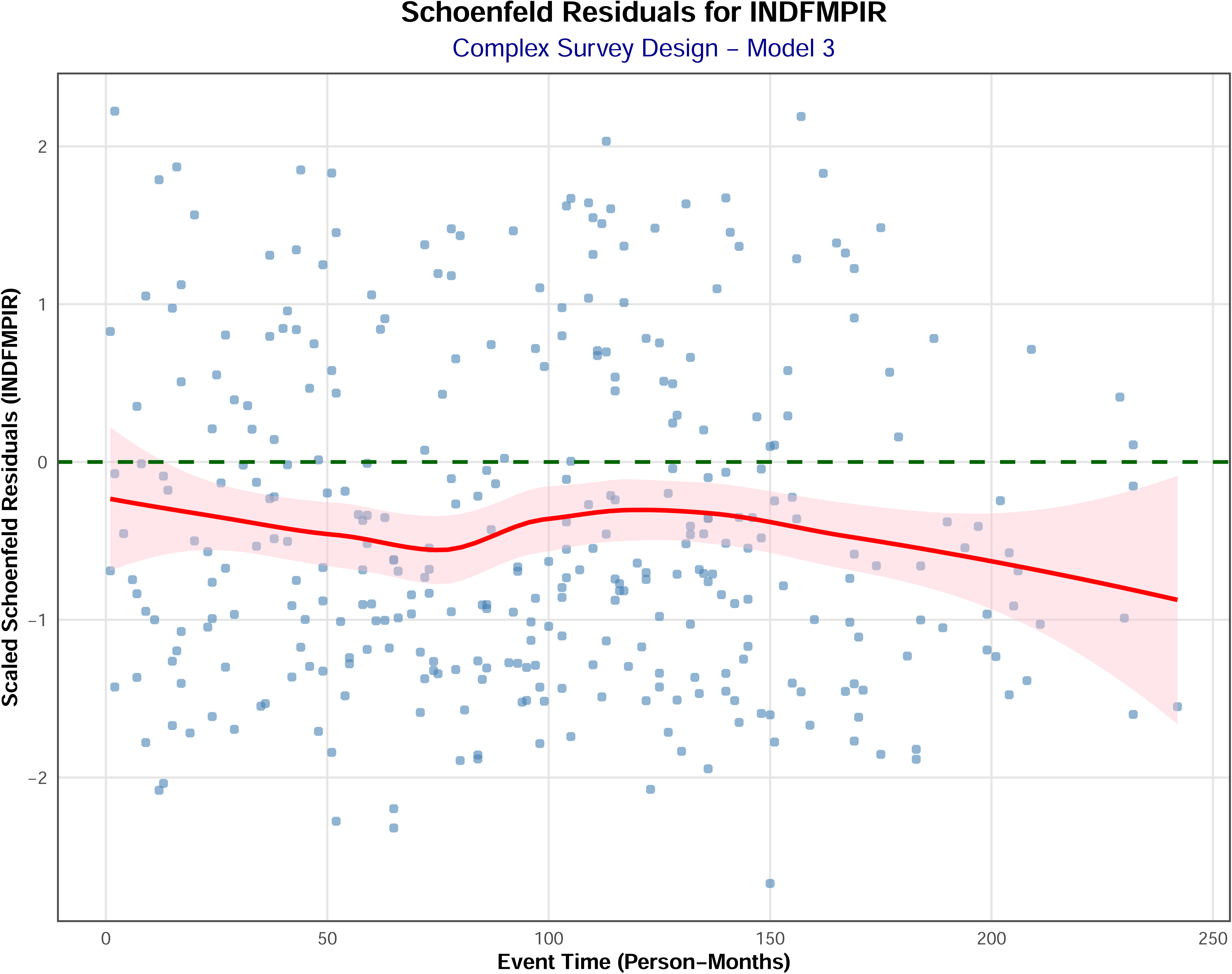

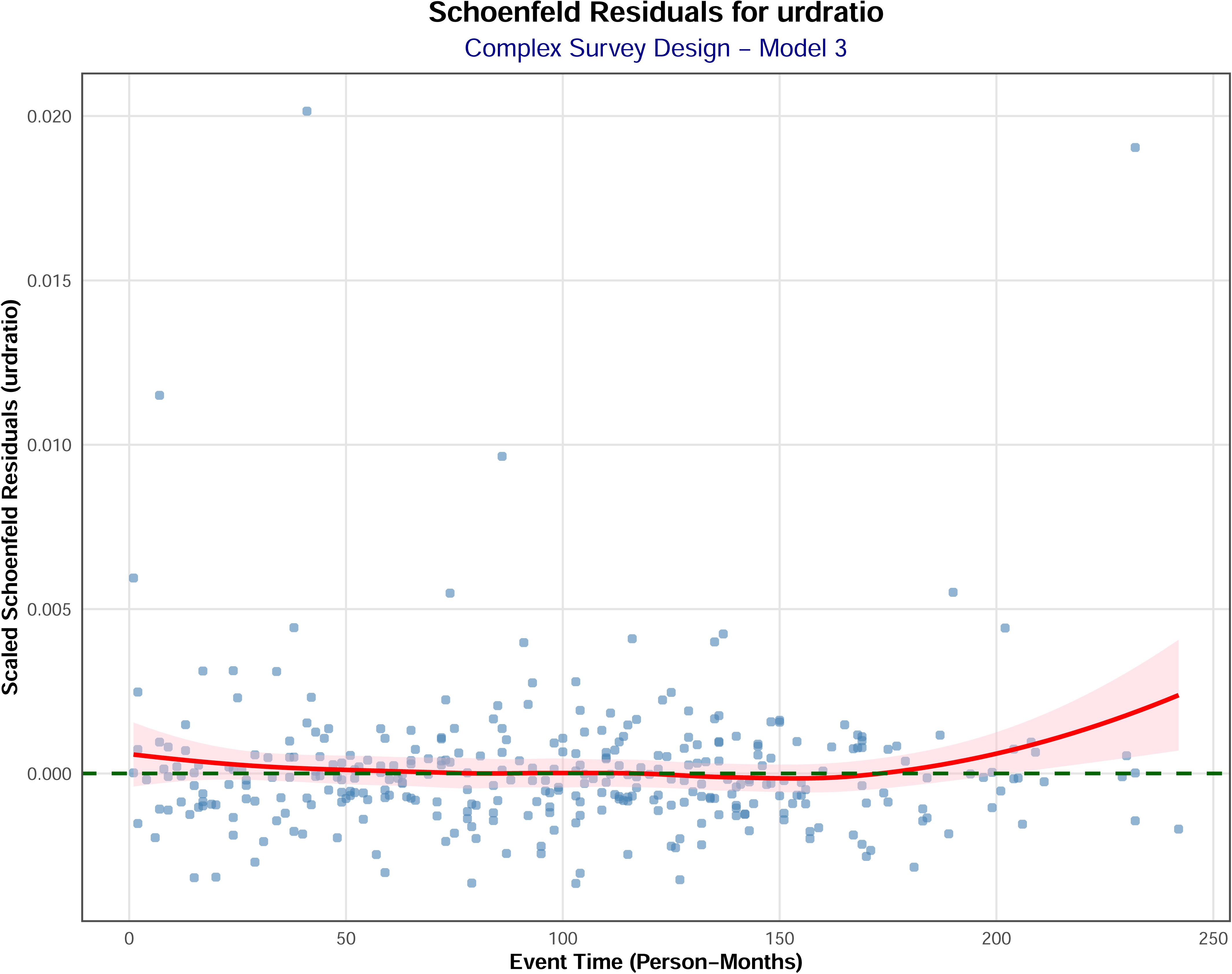

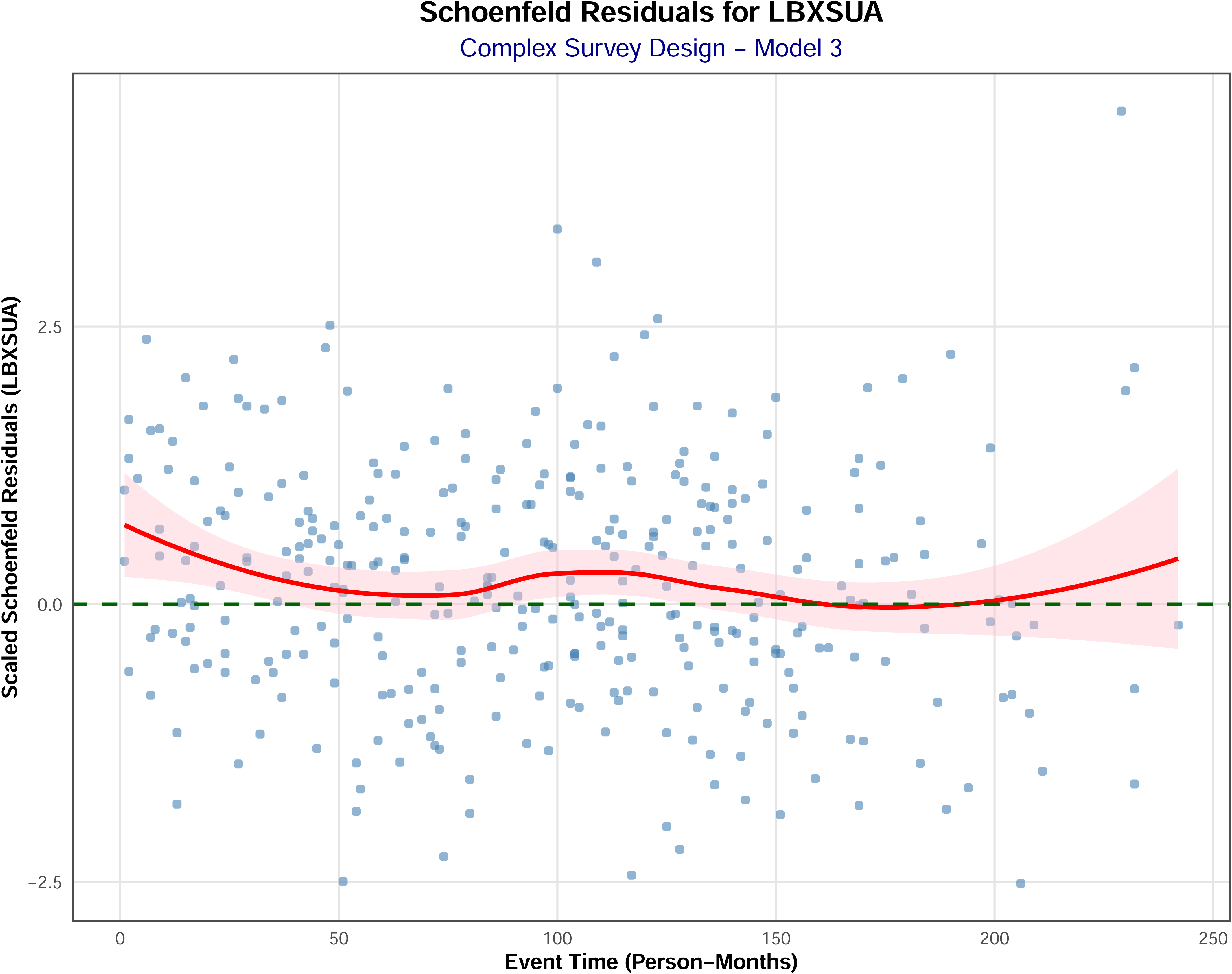

